# Host control of latent Epstein-Barr virus infection

**DOI:** 10.1101/2025.07.19.25331823

**Authors:** Axel Schmidt, T. Madhusankha Alawathurage, Leonard Frach, Sylvia Richter, Friederike S. David, Merle Schäfer, Sabrina K. Henne, Kaan Boztug, Anne-Katrin Pröbstel, Markus M. Nöthen, Eva C. Beins, Kerstin U. Ludwig

**Affiliations:** Institute of Human Genetics, University of Bonn, School of Medicine & University Hospital Bonn, Bonn, Germany; Department of Clinical, Educational and Health Psychology, Division of Psychology and Language Sciences, Faculty of Brain Sciences, University College London, London, UK; Clinic for Pediatric Immunology and Rheumatology, Center for Pediatrics and Adolescent Medicine, University Hospital Bonn, Germany; Center of Neurology, Department of Neuroimmunology, University Hospital & University Bonn, Bonn, Germany; German Center for Neurodegenerative Diseases (DZNE), Bonn, Germany; Departments of Biomedicine and Clinical Research and Research Center for Clinical Neuroimmunology and Neuroscience Basel (RC2NB), University Hospital Basel and University of Basel, Basel, Switzerland

## Abstract

Epstein-Barr virus (EBV) is a herpes virus that infects around 90-95% of the global population, and is associated with numerous autoimmune and neoplastic diseases. EBV persists in B cells as a life-long latent infection. Despite its relevance to disease development, the biological basis of host control during EBV latent infection remains unknown. Here, we report the identification of genetic and non-genetic factors that contribute to latent EBV infection control. In the genome sequence (GS) data of blood from 486,315 UK Biobank and 336,123 All of Us participants, we identified short-reads mapping to the EBV genome in 16.2% and 21.8% of individuals, respectively. The detection of EBV-reads (EBVread+) reflected increased viral load, and was associated with HIV infection, intake of immunosuppressive drugs, and current, but not former, smoking. Genome-wide association analyses identified strong associations at the Major Histocompatibility Complex (MHC), including 54 independent HLA-alleles of MHC class I and II, and at 27 genomic regions outside MHC (e.g., loci encompassing *ERAP2*, *CTLA4*). Associated genes included genes underlying monogenic susceptibility to EBV infection (e.g., *CD70*), and novel candidates thereof (e.g., *CD226*). In an analysis of individuals with EBV-associated diseases, we observed a higher polygenic burden of EBVread+ for HLA-alleles at MHC class I in multiple sclerosis (driven by HLA-A*02:01), and at MHC class II in rheumatoid arthritis (RA). A phenome-wide association analysis identified a polygenic overlap of EBVread+ with inflammatory bowel disease, hypothyroidism, and type I diabetes (T1D). Mendelian randomization analyses suggested potential causal effects of EBVread+ on RA and T1D. Our results will inform mechanistic studies and therapeutic approaches in EBV-associated diseases. More broadly, our study illustrates how by-products of human GS data can be used to investigate persistent viral infections.

## Introduction

Epstein-Barr virus (EBV; human herpesvirus 4) is a DNA-virus that infects ≈90-95% of the global population^1,2^. Primary EBV-infection usually occurs in childhood, when it remains asymptomatic or mild. If primary infection occurs during adolescence or beyond, EBV can cause infectious mononucleosis (IM)^5^. After primary infection, EBV persists mainly in memory B cells as a life-long latent infection^6,7^. Prior EBV infection is a risk factor for various cancers (e.g., Hodgkin-lymphoma (HL)), and autoimmune diseases (e.g., multiple sclerosis (MS), rheumatoid arthritis (RA), and systemic lupus erythematosus (SLE))^3,8,9^.

Inefficient control of latent EBV-infection may contribute to the development of EBV-associated diseases. While EBV is a prerequisite for MS^10^, only a subset of EBV-infected individuals develop the disease, following a prodromal phase of several years^11^. Furthermore, although MS risk is significantly elevated in individuals with prior IM, many MS patients did not have a severe primary EBV-infection^12^. Thus, MS may arise secondary to inefficient immune control of EBV during the prodromal phase, as indicated by high viral load^11^, potentially through molecular mimicry or chronic inflammation^9^. A plausible hypothesis is that similar mechanisms might be implicated in other EBV-associated autoimmune disorders, as supported by observations of elevated EBV viral loads in patients with SLE^13^ and RA^14^. In EBV-associated cancers, the importance of proper EBV immune control has been demonstrated by the analysis of inborn errors of immunity (IEI). IEI patients with impaired cytotoxicity of T and Natural Killer (NK) cells have an elevated EBV viral load in blood^15^, and are at increased risk for B cell derived EBV-positive lymphomas, such as HL, Burkitt lymphoma, or Diffuse Large B cell lymphoma^16^. Impaired EBV control also occurs in individuals with human immunodeficiency virus (HIV), or immunosuppression^17^. These groups also show an increased incidence of EBV-positive lymphomas^18,19^.

Despite its presumed relevance, few data are available concerning immune control during latent EBV-infection, which is largely due to a lack of appropriate data. While biobanks comprising hundreds of thousands of individuals with genetic data and health records are available, e.g., UK Biobank (UKB)^20^ and All of Us (AoU)^21^, these lack direct measures of EBV viral load. Within a subset of UKB, serology data on EBV are available. However, while these data can be used to identify prior EBV-infection, the role of serological factors in controlling latent EBV infection is limited^22^. Together, this prevents investigations into biological mechanisms and epidemiological correlations of latent EBV-infections.

To overcome these limitations, we exploited the fact that EBV-DNA is sequenced as a by-product of genome sequencing (GS) of human peripheral blood. Using GS data of peripheral blood from 486,315 UKB and 336,123 AoU participants, we demonstrated that the detection of short-reads mapping to the EBV genome (EBV-reads), is a robust surrogate measure for increased EBV viral load. We then investigated non-genetic and genetic influencing factors. At the non-genetic level, we demonstrated that the prevalence of EBV-reads was increased in immunosuppressed individuals, in current, but not former smokers, and in samples obtained in winter. At the genetic level, we found a strong association of the MHC locus and of 27 loci outside of the MHC, which were broadly consistent across the two biobanks, and suggest novel candidate genes for key roles in EBV immunity. Analyses of the genetic overlap with EBV-associated diseases generated novel hypotheses regarding mechanisms in MS and RA, and identified novel diseases for which EBV might be pathophysiologically relevant, e.g., type 1 diabetes (T1D).

## Results

### Detection of EBV-reads using GS data from UKB and AoU

We retrieved short reads mapping to EBV (EBV-reads) from GS data of 490,294 UKB participants (Methods, Supplementary Notes). During quality control (QC), 51 library-preparation plates showed an unexpectedly high number of individuals with EBV-reads (**Supplementary Fig. 1**). Since this might indicate contamination, these plates were excluded. The aggregated EBV-reads of the remaining 486,315 individuals (*UKB-QC-*cohort, **Fig. 1a**) showed uniform EBV-genome coverage (NC_007605.1, **Fig. 1b**). The EBV-read distribution was zero-inflated, with most individuals showing no EBV-reads (n=407,544; 83.8%, denoted as “EBVread-”; **Fig. 1c**). Of the 78,771 individuals in whom EBV-reads were detected (“EBVread+”, 16.2%), 61.9% had exactly one read (maximum: 27,639 reads in one individual). EBV-reads were also extracted from the GS data of 365,931 AoU participants (Methods). Following QC, the AoU dataset comprised 336,123 individuals (*AoU-QC*-cohort). The EBV-read count distribution was similar to that observed in UKB (**Supplementary Fig. 2**). Overall, 21.8% of the participants (and 17.6% among the European subgroup) were EBVread+ (**Supplementary Table S1**). The fraction of EBVread+ individuals identified from GS in UKB and AoU are consistent with smaller studies of immunocompetent individuals, as based on GS data (14.0%)^23^ or quantitative PCR (11.03%)^18^.

**Figure 1:**
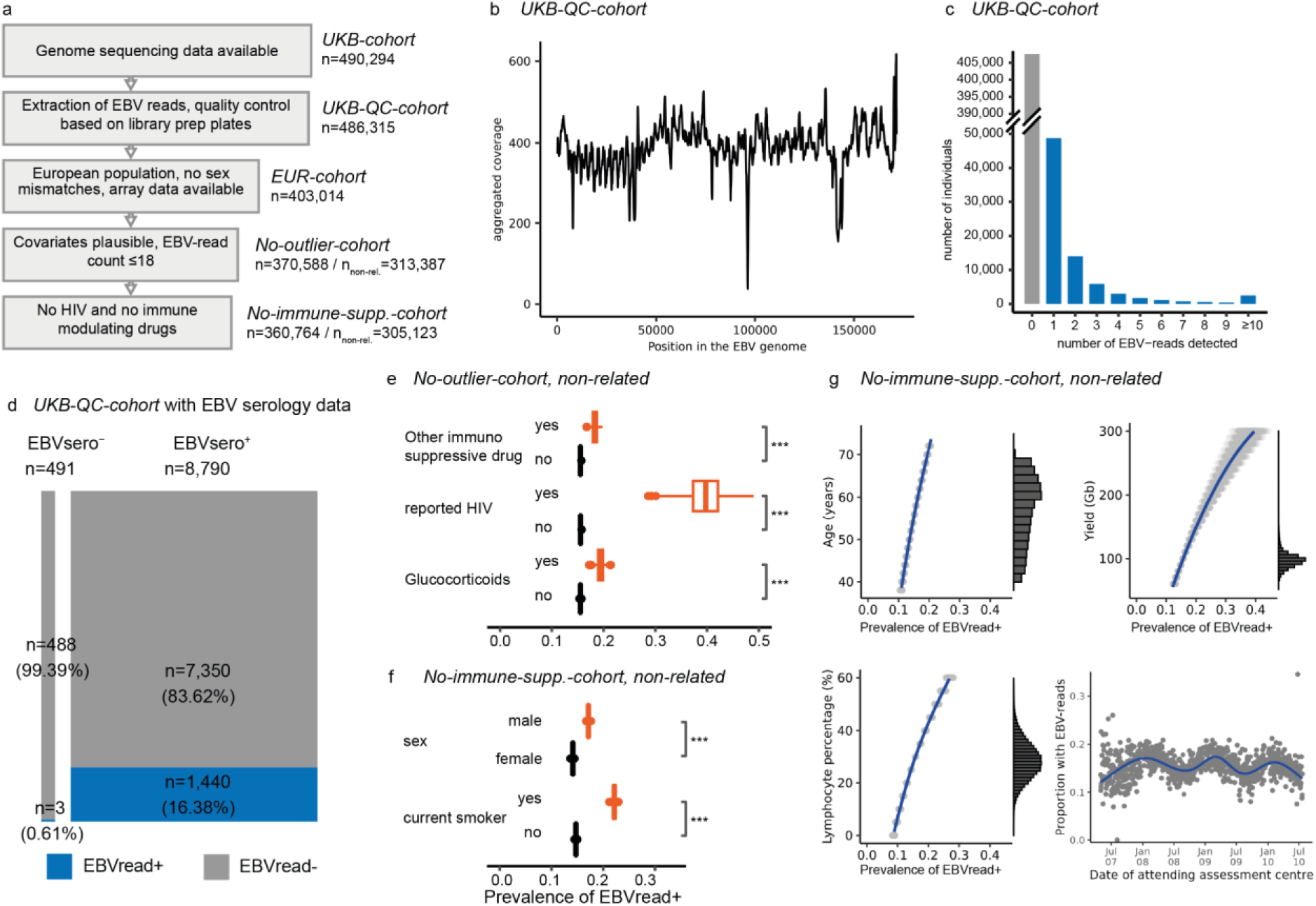
Analysis of EBV-read abundance in blood-based genome sequencing data of UK Biobank. **a)** Flowchart of cohort definitions created within UK Biobank (UKB), as based on consecutively applied parameters. Number of individuals (n) refers to related individuals, unless otherwise indicated (non-related (n_non-rel._)). **b)** Cumulative read coverage across the EBV-genome using the UKB-QC-cohort (line smoothed, rolling windows of 500 bp). **c)** Number of EBV-reads detected per individual is zero-inflated. Individuals with at least one EBV-read are highlighted in blue (EBVread+). Note that the y-axis is interrupted between 50,000 and 390,000 individuals. **d)** In a subcohort with available EBV serology data (EBVsero-negative (EBVsero-), EBVsero-positive (EBVsero+)), EBVread+ status was highly specific, but not sensitive, for being EBVsero+. **e-g)** Several non-genetic factors were associated with the probability of detecting EBV-reads: In the *no-outlier*-cohort, immune-modulating factors significantly increased the prevalence of EBVread+ individuals (panel e). Following the exclusion of immunocompromised individuals (Methods), male sex and current smoking status were both associated with an increased probability of EBV-read detection (f). Additionally, higher age, lymphocyte percentage, sequencing yield, and sampling date in winter season were associated with increased probabilities of detecting EBV-reads. (e-g). Estimates were obtained using marginalization, and distributions were estimated using 1000 bootstrap replicates (Methods); ***: consistent across 1000 bootstrap replicates.

Next, we derived a subcohort of UKB individuals with available serology data (“serology cohort”, n=9,281). Based on prior definitions^1^, 491 individuals were considered EBVseron-egative (sero-) and 8,790 EBVsero-positive (sero+; Methods). A comparison of EBVread+/- and EBVsero+/- status revealed that while being EBVread+ was highly specific, it was not a sensitive indicator of EBVsero+ status (specificity: 99.4%, sensitivity: 16.4%; **Fig. 1d**). To investigate whether EBVread+ is a measure of elevated viral load, we simulated the sequencing process under the assumption that EBV viral load in blood cells follows a log-normal distribution, as reported for HIV-1 viral load^24^ (**Extended Data Fig. 1**). The distribution of detected EBV-read counts was compatible with a log-normal distribution of the underlying EBV viral load, suggesting that the analysis of EBVread+ versus EBVread-represents an approximation of EBV viral load within human blood samples.

### Non-genetic factors contributing to EBVread+

The influence of non-genetic factors on EBVread+ was investigated in order to: (i) identify such factors; (ii) exclude from further analysis all individuals whose EBV count was essentially determined by a strong exogenous factor; and (iii) control for these factors in further analyses. Herefore, we first, we examined associations between 11,111 SNOMED concept IDs and EBVread+/- in AoU (Methods). Initial test statistics were highly inflated, with HIV-positivity and smoking showing the strongest associations. When the analysis was conditioned on these two traits, inflation was largely resolved (**Supplementary Table S2**). Nonetheless, associations between the probability of being EBVread+ and several immune-related SNOMED concepts remained (**Supplementary Fig. 3**). In the top EBV-read count percentile, we observed a high prevalence of individuals with pathophysiological processes that cause an increased EBV viral load (e.g., neoplastic diseases, pronounced immunosuppression; **Supplementary Fig. 4**).

We then aimed to: (i) quantify the effect of HIV and immunosuppression on EBVread+; and (ii) identify additional contributors. To avoid overfitting, we used the independent *UKB-QC-*cohort, and increased its homogeneity by: (i) excluding individuals who were in the top EBV-read count percentile or had outlier blood count measurements; and (ii) limiting the analysis to non-related individuals of European ancestry (“*no-outlier*-cohort”; **Fig. 1a**, Methods). Of these 313,387 individuals, 48,771 (i.e., 15.6%; with an expected standard deviation (s.d.) of 0.1% based on bootstrapping) were EBVread+. Both HIV-infection and immune-modulatory drugs significantly increased the likelihood of being EBVread+. The highest probability was found for reported HIV infection (39.7%, s.d.=3.5%; **Fig. 1e**), followed by intake of glucocorticoids (19.4%; s.d.=0.7%) or other immunosuppressive drugs (18.3%; s.d.=0.5%).

To identify further contributing factors in immunocompetent individuals, we excluded from the *no-outlier* cohort individuals with reported HIV infection or use of glucocorticoids or other immunosuppressive drugs (“*no-immune-supp-*cohort”, n=305,123), and performed variable selection on a set of predefined covariates (47,234 EBVread+, 257,899 EBVread-; Methods, **Supplementary Table S3**). EBV-reads were detected more frequently in males compared to females (17.1%; s.d.=0.1% vs. 14.1%; s.d.=0.1%), and in current-smokers compared to current non-smokers (22.1%; s.d.=0.3% vs. 14.7%; s.d.=0.1%; **Fig. 1f**). Former smoking status was not selected by variable selection, suggesting that the lower prevalence of EBVread+ in current non-smokers is driven by both former and never smokers. In contrast, increasing age, GS yield, and lymphocyte percentage were selected, and were positively correlated with the presence of EBV-reads (**Fig. 1g**). A seasonality effect was also observed, i.e., EBV-read detection was more probable in winter than in summer (**Fig. 1g**). This observation was independently confirmed in AoU (**Supplementary Fig. 2**).

### Identification of common genetic variants associated with EBVread+

To identify common genetic variants associated with EBVread+, a genome-wide association study (GWAS) was performed using the “*no-immune-supp* cohort” and imputed data (n=56,180 EBVread+ cases, n=304,103 EBVread-controls; Methods). Variants at 28 loci showed genome-wide significance, including an extended association at the MHC locus and additional associations at 27 non-MHC loci (**Fig. 2a**, Methods, **Supplementary Fig. 5**, **Supplementary Table S4**).

**Figure 2:**
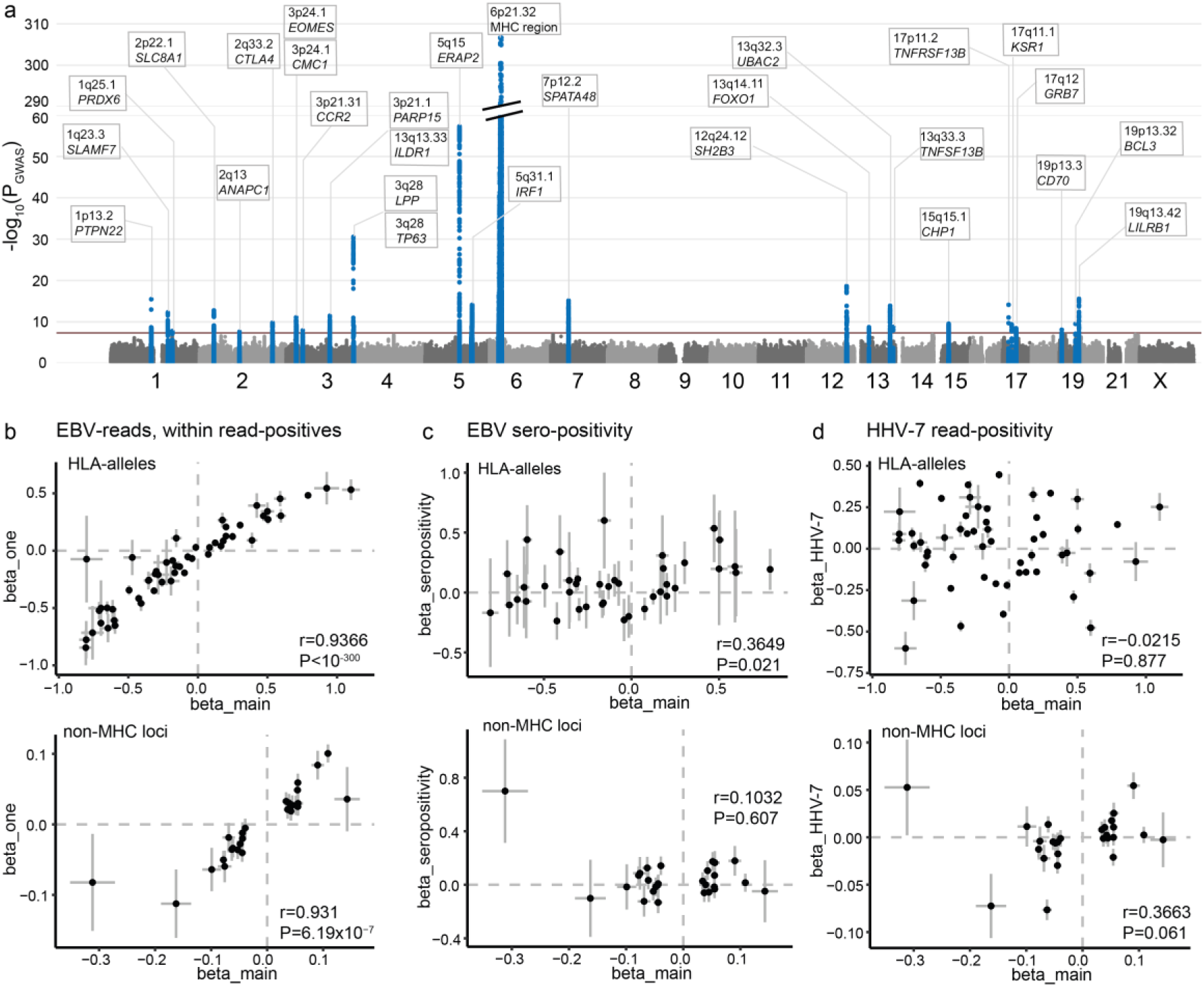
Genetic association analyses and correlation of effect sizes. **a)** Manhattan plot of a common variant-based genome-wide association study (GWAS) using 56,180 EBVread+ cases and n=304,103 EBVread-controls from the no-immune-supp cohort (excluding individuals without imputed genotype data). Significant loci (P<5×10^-8^ red line) are indicated in blue. Note that the y-axis showing the negative decadic logarithm is interrupted between 60 and 290. **b)** Effect sizes of independently significant HLA-alleles (top row) and lead variants at 27 non-MHC loci (bottom row; P<5×10^-8^) from the EBVread+ GWAS (beta_main) are plotted against effect sizes from a within-EBVread+ GWAS (controls: one EBV-read, cases: higher EBV-reads). Spearman correlation coefficients (r) and respective P-values (P) are indicated in the right-bottom corner. Note that the P-value for HLA-alleles was too small for reliable calculation. For **c)** and **d)** the plots are similar to those presented in c); except for EBV seropositivity (c) and the presence of HHV-7 reads (d). Seropositivity (in c) was defined as having at least two out of four antibody titers exceeding predetermined thresholds^29^. Error bars indicate standard error.

At the MHC region, the immunologically relevant variants are alleles of HLA-genes („HLA-alleles“) which determine the antigen repertoire that individuals can present to the adaptive immune system. We attributed the extended MHC association to 116 different HLA-alleles (**Supplementary Table S5**), which were reduced to 54 independent alleles after iterative conditional analyses (at P<5×10^-8^; **Supplementary Table S6**, Methods). Notably, these included alleles from both MHC class I and MHC class II. The lowest P-value was observed for MHC class II allele HLA-DRB1*04:04, which had a substantial effect size for EBVread+ (beta=0.79; s.e.m.=0.02) and has been associated with increased RA risk^25^. The second and third most significant HLA-alleles were HLA-A*02:01 (beta=-0.31; s.e.m.=0.01) and HLA-B*14:02 (beta=-0.68; s.e.m.=0.02). Both alleles were associated with a reduced likelihood of EBVread+, and HLA-A*02:01 has been reported to decrease the risk for MS^26^, EBV-positive HL^27^, and endemic Burkitt’s lymphoma^28^.

To ensure the robustness of our results, we performed complementary analyses. The effect sizes of the EBVread+ GWAS lead variants at the 27 non-MHC loci and the 54 independent HLA-alleles were compared to effect sizes from complementary association analyses, using Spearman correlation (**Supplementary Table S7**). First, we calculated effect sizes for a modified case-control definition in UKB that better reflects EBV viral load, though at lower sample sizes (EBV-read count = 1 (n=35,703) vs. EBV-read count ≥ 2 (n = 20,477)). We observed highly significant correlations of effect sizes for both variant sets (non-MHC loci: r = 0.93, P = 6.2×10^−7^; HLA-alleles: r = 0.94, P < 10^-300^; **Fig. 2b**). In contrast, only weak evidence was generated for correlations of effect sizes to EBV sero-positivity (**Fig. 2c**)^29^, although we observed some suggestive correlations with different antibody levels for the HLA-alleles (Methods, **Extended Data Fig. 2**).

Since memory B cells are the main latency reservoir for EBV^4^, their variability in blood samples could confound EBVread+ associations. Using a recent GWAS of memory B cell abundance^30^, we found no significant overall correlation of effect sizes at non-MHC loci (**Extended Data Fig. 2**, no MHC-data provided). However, one locus was associated with both traits (13q33.3_*TNFSF13B*, **Supplementary Table S8**). To investigate the specificity of the EBVread+ GWAS loci, we next performed association analyses of human herpesvirus 7 (HHV-7) abundance in GS-data from UKB (Methods; **Supplementary Fig. 6**). We found no significant correlation for HLA-alleles and non-MHC risk loci. However, a trend towards positive correlation for non-MHC risk loci was observed (r = 0.37, P = 0.06; **Fig. 2d**). Six of the non-MHC loci had *P*<0.05, with a consistent effect direction. For two of these, shared causal variants were observed, as based on colocalization analyses (*SLC8A1, PTPN22;* posterior probability (H4)>0.5, **Supplementary Table S9**). These findings suggest that the genetic associations with EBVread+ represent specific factors associated with EBV viral load during latency. They do not seem to be confounded by memory B cell abundance, with the exception of the *TNFSF13B* locus, which has an established role in memory B cell survival^31^.

Finally, we aimed to replicate our EBVread+ GWAS results in 184,948 individuals of European population background from the *AoU-no-outlier* cohort. Of the 116 associated HLA-alleles, 106 were matched to HLA-alleles in AoU. For 98 of these, nominal significance and a consistent effect direction were observed (including 77 HLA-alleles with genome-wide significance in AoU; **Supplementary Table S5**). Lead variants at 25 of the 27 non-MHC loci were also replicated (at P<0.05; **Supplementary Table S4**).

### Annotation and fine-mapping of associated non-MHC loci

We then characterized the associations at the 27 genome-wide significant non-MHC loci, based on the respective lead variants. Similar to most multifactorial traits, the majority of lead variants mapped to non-coding regions. We applied four complementary approaches to identify potential effector genes at these loci, and highlight 22 genes at 17 loci which were implicated by at least three approaches (Methods, **Supplementary Table S9**). These included genes with an established role in immune processes (e.g., *ERAP2*, *EOMES*), and genes underlying known IEIs (e.g., *CTLA4*).

To identify target genes that might be missed by the four position-based approaches, we retrieved cell type-specific cis-eQTL data generated in peripheral blood mononuclear cells (PBMC) from the OneK1K project^32^ (Methods). A total of 18 variant-gene-cell type associations were identified for EBVread+ lead variants or proxies thereof (single-nucleotide polymorphisms (SNPs), R^2^>0.7), most of which concerned *ERAP2* across multiple cell types (**Supplementary Table S10**). *ERAP2* encodes an aminopeptidase which trims peptides within the endoplasmic reticulum prior to loading onto MHC class I^33^. Three common haplotypes are present at the *ERAP2* locus, two of which (B and C) harbor established splice variants that cause *ERAP2*-mRNA to undergo nonsense mediated mRNA decay (NMD), rendering them non-functional^33^. Both NMD haplotypes are tagged by the G allele of rs2927608. Besides being associated with reduced *ERAP2* expression in eQTL data, this eSNP-allele was associated with a higher prevalence of EBVread+ in our GWAS (**Fig. 3a**). Other cell type-specific eQTL effects were found for *CTLA4* and *CMC1* in S100B positive CD8+ T cells, and *SLC22A5* in NK cells (**Supplementary Table S10**).

**Figure 3:**
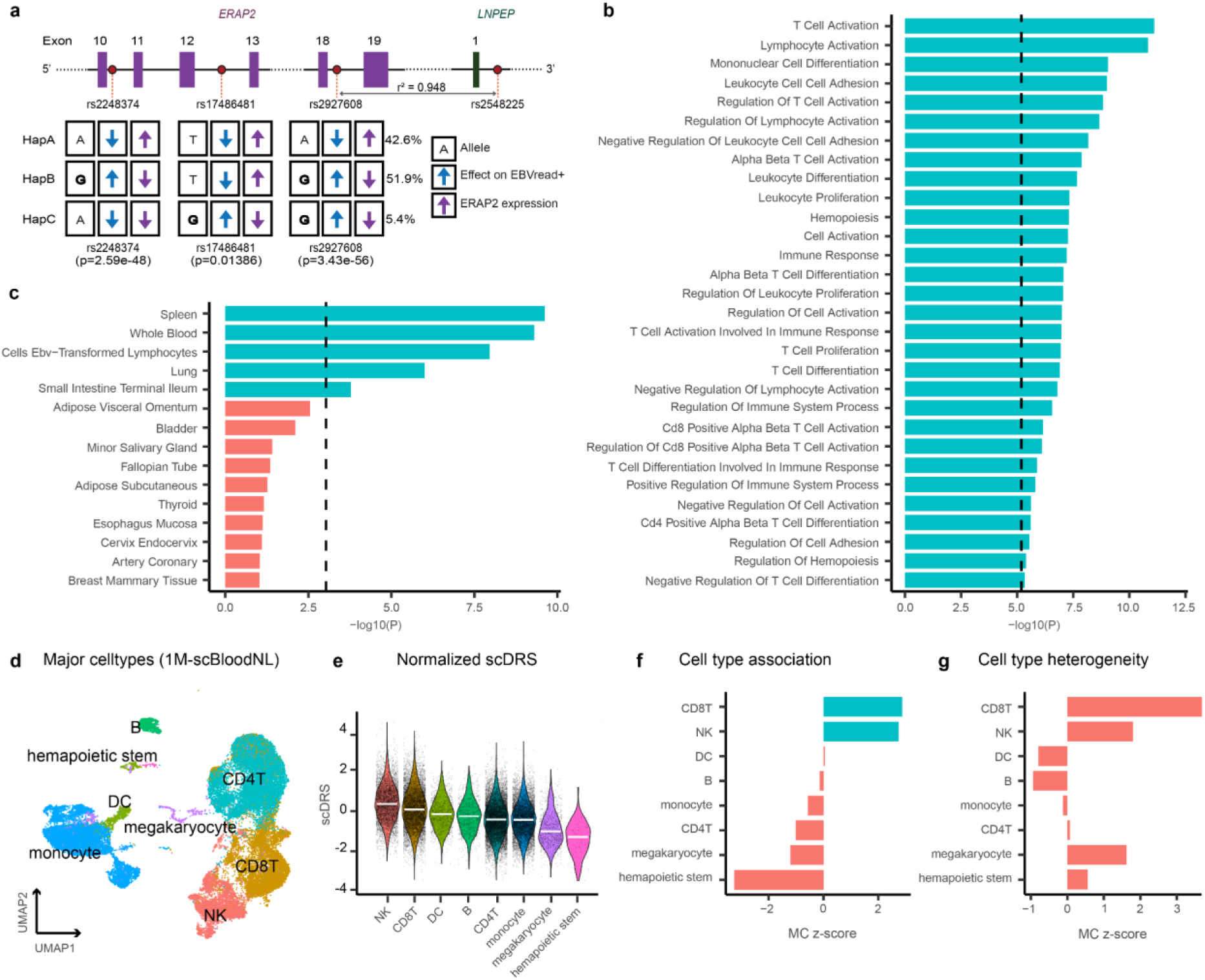
Characterization of non-MHC risk loci associated with EBVread+. **a)** Schematic representation of associated variants at the *ERAP2* locus (statistical lead variant: rs2548225). *ERAP2* exons (starting from exon 10) are shown with the location of splice site variants that define different *ERAP2* haplotypes (HapA–C). HapB and HapC encode mRNAs that undergo nonsense-mediated decay, and are associated with reduced expression based on GTEx v8 (whole blood). Variant rs2927608 is an eSNP for *ERAP2* in the OneK1K data^32^. For each allele, the direction of effect on *ERAP2* expression and EBVread+ association is indicated by an arrow. Risk alleles associated with EBVread+ are highlighted in bold. **b)** Bar plot of –log₁₀ *P*-values for Gene Ontology Biological Process (GOBP) terms that are significantly enriched for genes with significant associations for common variants. Only GOBP exceeding the Bonferroni threshold of P=6.5×10^-6^ (dashed line) are shown. **c)** Similar analyses and plot as in b), using gene expression levels across 54 GTEx tissues (dashed line: Bonferroni correction threshold of P=9.2×10^-4^). The top 15 most significant GTEx tissues are shown, with blue color indicating significant GTEx tissues. **d)** UMAP representation plot of the PBMC single-cell RNA sequencing (scRNA-seq) data^40^, colored by cluster labels of cell type annotation level 1. **e)** Distribution of normalized single-cell disease relevance scores (scDRS) across cell types of annotation level 1, presented in descending order based on median scDRS (white bar). Higher scores indicate cells with excess expression of genes implicated by the EBVread+ GWAS. Results of the Monte Carlo (MC)-based statistical inference of (**f**) cell type association and (**g**) within-cell type heterogeneity with scDRS based on EBVread+. Bars are colored based on significance: blue indicates a multiple comparison-adjusted false discovery rate (FDR) **<**0.05, red indicates non-significant values.

To investigate potential pleiotropic effects, associated phenotypes for lead variants at the 27 non-MHC loci were retrieved from OpenTargets (Methods). Among a total of 3,745 associations (at P<0.005), the largest overlap was found for diverse blood count measures, followed by asthma (9 loci), T1D (7 loci) and myocardial infarction / SLE (6 loci respectively; **Supplementary Table S11**). In addition, we observed high pleiotropy for some loci (>100 associated phenotypes, e.g., loci including *SH2B3*, *PTPN22,* and *IRF1*), while other lead variants had only a few associations at the same significance threshold, potentially indicating a specific role in EBV control (e.g., *ILDR1*, *CMC1*). For some of the prioritized genes at associated loci, we found evidence for pharmacological relevance (**Supplementary Table S12**). For example, EBVread+ associated variant rs66692283 at 1q23.2 is an eQTL for *SLAMF7*, with the risk allele increasing *SLAMF7* expression, suggesting a potential therapeutic target for the inhibitor Elotuzumab.

To identify putative causal variants at individual non-MHC loci, we applied statistical fine-mapping and identified 28 credible sets of variants (CS; posterior inclusion probability (PIP) >95%) at 23 loci (**Supplementary Table S4**). Of these 889 CS variants, three were missense variants with individual PIPs >0.1 (p.Trp620Arg in *PTPN22* (PIP: 0.51), p.Trp262Arg in *SH2B3* (0.19), and p.Cys104Arg in *TNFRSF13B* (1.0), **Supplementary Table S13**). These might represent the functionally relevant variants at these loci. The CS also contained one non-coding variant at PIP >0.95, i.e., rs531660643. This is a reported splice QTL for *BCL3* in whole blood (**Supplementary Fig. 7**), whose gene product BCL3 is involved in B cell fate and regulates NFκB^34^. Using SNPs outside the MHC region, we estimated a SNP-based heritability of 2.04% (s.e.m.=0.44%; LD Score regression^35^) for EBVread+.

### Gene-based analyses suggest novel genes for involvement in host-EBV interaction

After analyzing individual loci, we applied genome-wide approaches to: (i) identify novel genes for immune control of EBV; and (ii) enable systematic analyses on the biology of host-EBV interaction.

First, gene-based association statistics were calculated using common variants from our EBVread+ GWAS and MAGMA^36^ (without MHC-region). We identified 63 significant genes after Bonferroni correction, the majority of which are located at the 27 non-MHC risk loci (described above). We found 10 additional genes at five loci, which had failed to reach genome-wide significance in the single-variant analysis (e.g., *NFKB1*). Among 2,083 genes with significance at P<0.05, we observed 83 genes classified as IEI, including genes underlying monogenic EBV-driven lymphoproliferative disease (*RASGRP1*, *CORO1A*, *SH2D1A* and *CD70*), and broader IEIs (e.g., *NFKB1*, *IKZF1*, *IKZF3*; **Supplementary Table S14**). To formally test for an enrichment, we applied MAGMA gene-set enrichment. We found that genes causing IEIs (n=456) showed highly significant enrichment for association with EBVread+ (*P*=4.66×10^-6^, beta=0.19; s.e.m.=0.04). When considering a subset of 14 genes that specifically cause monogenic EBV-driven lymphoproliferative diseases^15^, the effect size further increased (beta=0.35; s.e.m.=0.22, *P*=0.055; **Supplementary Table S15**).

Gene-based association analyses of rare variants (RVAS_gene_) with minor allele frequency (MAF) <1% were then performed, using individuals from the *no-immune-supp.-*cohort with available exome sequencing data (n=54,259 EBVread+, n=293,834 EBVread-). Based on four variant pathogenicity definitions (“masks”; see Methods) and 18,796 protein-coding genes, 29 genes showed at least one test-wide significant mask (P_gene_< 8.86×10^-7^, n=55 associations, **Supplementary Table S16**). Twenty-eight of these genes were located within the MHC locus. The only non-MHC gene was*TNFRSF13B*, whose signal was driven by a single variant (p.Cys104Arg), as demonstrated by a non-significant result when this variant was excluded (P = 0.087). This variant is associated with the development and clinical course of common variable immunodeficiency (e.g., autoimmunity, lymphoproliferation)^37^, and with tonsillectomy and ear surgery in the general population^38^. Overall, we observed 22 IEI genes at a more lenient threshold (P<0.01), including *TAP1* and *TAP2* at the MHC region, but also *ITK,* which causes a monogenic immune deficiency in which EBV control is specifically impaired^15^.

When comparing the overlap of genes enriched in both GWAS-based MAGMA and RVAS (at P<0.01, excluding MHC-region), 24 genes had complementary evidence from common and rare coding variants for a role in host-EBV control (**Supplementary Table S17**). These included seven genes whose rare variant enrichment was driven by putative loss-of-function variants (*PTPN22*, *GP1BA*, *CD226*, *C6orf222*, *ZNF284*, *CHD4*, *HKR1*), representing strong novel candidate genes for host control of latent EBV-infection.

### Identification of candidate pathways and effector cell types

To generate insights into effector pathways, tissues, and cell types, we used gene-based association statistics from MAGMA and performet gene-set enrichment analyses^39^. First, we used gene-sets corresponding to Gene Ontology Biological Processes (GOBP), and identified 30 test-wide significant pathways (**Fig. 3b, Supplementary Table S18**). These encompassed various immune pathways, e.g., T cell activation and differentiation, thus supporting the known role of T cells in latent EBV control. Second, we used expression information from 54 tissues available in GTEx v8, five of which (i.e., spleen, whole blood, EBV-transformed lymphocytes, lung, and terminal ileum) were identified as potential effector tissues (**Fig. 3c**). We hypothesized that tissue resident leukocytes might be partially responsible for the observed enrichments in spleen, lung, and terminal ileum, which must be demonstrated by analyses of single-cell data.

To disentangle the enrichment in blood or leukocyte-rich organs, we used a PBMC gene expression dataset^40^, and performed enrichment analyses of individual cell types using the single-cell disease relevance score approach^41^ (scDRS; Methods, **Fig 3d, e**). Analysis of MAGMA candidate genes in eight major cell types (annotation level 1) revealed significant enrichments in CD8+ T cells, consistent with their role in eliminating EBV-infected B cells^42^ and NK cells. At a more fine-grained annotation level (level 2, involving 21 cell types), the highest average scDRS was observed in the small cell cluster annotated as NK_bright_ cells. Further support was also generated for NK_dim_ and memory CD8+ T cells, both of which have similar enrichment P-values. albeit for much larger cell numbers (**Supplementary Fig. 8**).

### Polygenic architecture of EBVread+ and EBV-associated diseases

Next, we evaluated whether an aggregated genetic risk score (GRS) generated in individuals of European ancestry: (i) improves risk prediction for EBVread+; (ii) is transferable across cohorts and ancestries; (iii) is associated with known EBV-associated diseases; and (iv) can be used to identify novel EBV-candidate diseases through phenome-wide association studies (PheWAS). Herefore, we first created target disease cohorts by assigning affected individuals from the UKB *no-outlier* cohort to IM, HL, MS, RA, non-Hodgkin-Lymphoma (NHL), SLE, and Sjoegren disease (**Supplementary Table S19**). Individuals with EBV serology data were used as a control cohort, and individuals not included in any of these groups were used as base-cohort (**Fig. 4a**). In the base-cohort, we generated different GRS, based on imputed HLA-alleles or genotyped SNPs (Methods). In the control cohort, the GRS encompassing all HLA-alleles (HLA_all) best explained EBVread+ according to Nagelkerke R^2^ (improvement over the base-model: ΔR^2^ = 0.079 ± 0.009 s.d., **Fig. 4b**). To capture different components of biology, for the subsequent analyses we selected: (i) HLA_all; (ii) its derivatives HLA_MHC I and HLA_MHC II, which can be considered uncorrelated predictors; and (iii) a GRS based on all non-MHC SNPs (SNP_wo_MHC). All four GRS were positively correlated with increased EBV-read counts in the control cohort (**Fig. 4c**, **Extended Data Fig. 3**), and did not differ between EBVsero+ and EBVsero-groups (**Fig. 4d**).

**Figure 4:**
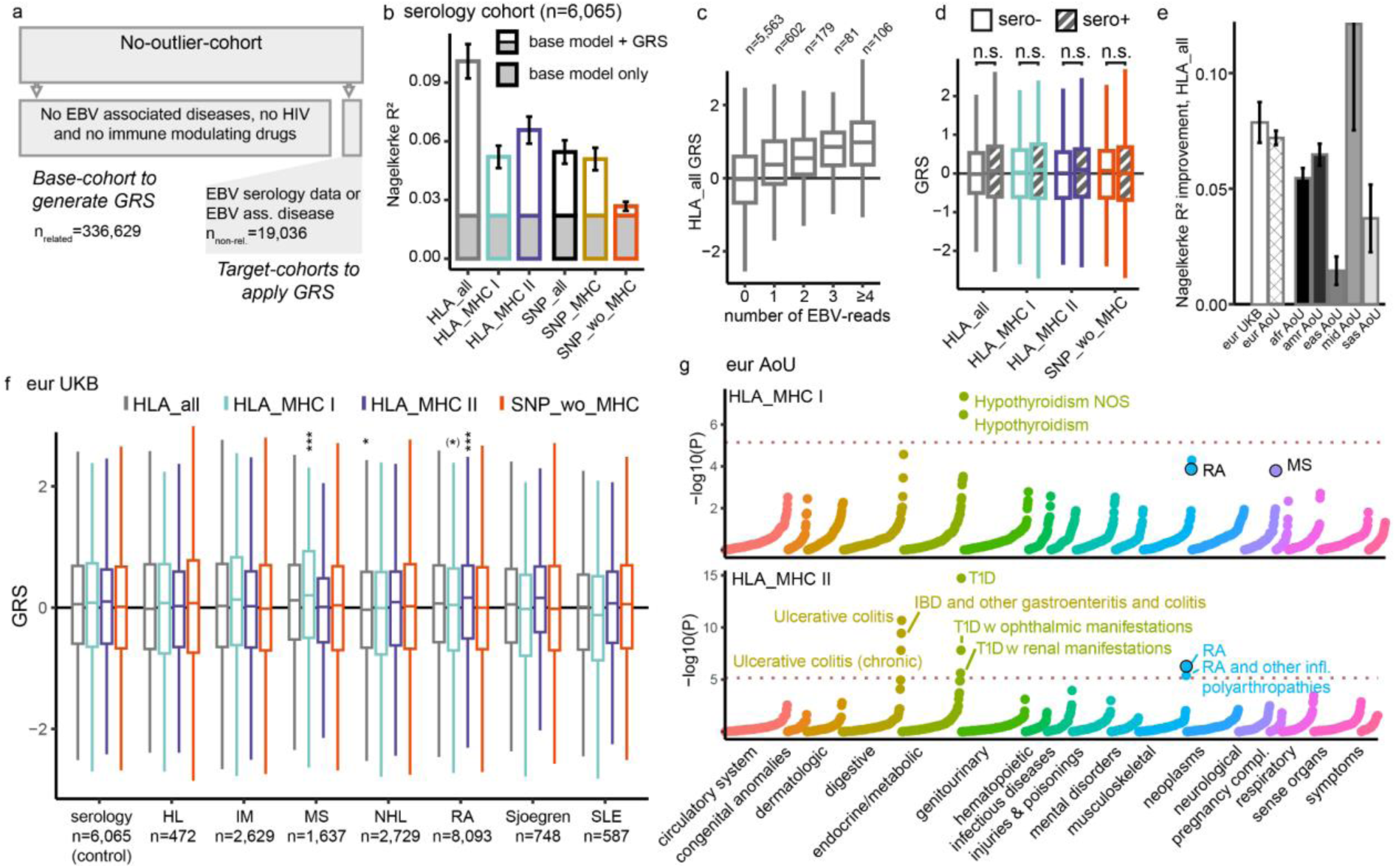
Genetic risk score analyses in UKB and AoU. **a)** To generate independent cohorts for the genetic risk score (GRS) analyses, the UKB *no-outlier-*cohort was divided into a discovery (base) cohort and additional target cohorts. The latter included (i) selected EBV-associated diseases and (i) a control cohort of individuals with serology data. **b)** In the serology cohort, EBVread+ status was first predicted using a base model (including age, sex). This analysis was then repeated by an analysis of the base model and one of six GRS: all imputed HLA-alleles (HLA_all); HLA-alleles of MHC I (HLA_MHC I); HLA-alleles of MHC II (HLA_MHC II); all genotyped SNPs (SNP_all); SNPs within the MHC locus only (SNP_MHC; position within 25-36Mb on chromosome 6, GRCh37); or all variants except variants within the MHC locus (SNP_wo_MHC). The respective values for Nagelkerke R^2^ were calculated and are plotted on the y-axis. **c)** The GRS for HLA_all, which was considered the best performing GRS according to b), was positively correlated with the number of observed EBV-reads. **d)** No significant difference was observed in GRS distribution between sero- and sero+ individuals in the serology cohort for any of the scores. This provides further evidence for the hypothesis that the genetic factors involved in controlling EBV viral load are different at least in part to those involved in sero-positivity. P-values were calculated using logistic regression, with adjustment for covariates and likelihood ratio tests (Methods). **e)** Improvements in Nagelkerke’s R^2^ relative to base models within UK Biobank (taken from b), and six continental populations in AoU for the HLA_all GRS (Methods). **f)** GRS distributions of individuals within the serology cohort and individuals with selected EBV-associated diseases. The black line corresponds to the mean GRS (all) of all cohorts. Note that each group includes a number of individuals with multiple diseases. Disease abbreviations as in the main text. P-values were calculated as described in d. **g)** Negative decadic logarithms of P-values of individual PheCodes, grouped according to organ systems or disease groups are displayed. The upper panel represents results for HLA_MHC I, the lower for HLA_MHC II. The dashed line corresponds to the Bonferroni corrected significance threshold, and phenotype names are given for significant results or for associations identified in f (highlighted in black). P-values were calculated as described in d. **c, d and f)** The elements of box plots correspond to the following values: thick line: median, box: 25th and 75th percentile, whiskers: largest / smallest value no further away from the box than 1.5 times the interquartile range. In g) p-values were Bonferroni-corrected. ***: P<0.001; *: P<0.05; ^(^*^)^: P<0.1.

Next, we applied these four GRS to the *AoU-no-outlier* cohort, which was split according to genetic ancestry (Methods). In the AoU European subcohort, which had the highest genetic similarity to the base-cohort, the improvements in Nagelkerke R^2^ values compared to the base model were similar to our results from UKB, with HLA_all best explaining EBVread+ (ΔR^2^ = 0.072; s.d. = 0.002; **Fig. 4e; Extended Data Fig. 3**). Similarly, HLA_all showed the largest improvements in Nagelkerke R^2^ in each of the five non-European ancestry groups, despite differences in the absolute values (**Fig. 4e)**. In individuals from the African (ΔR^2^ = 0.055; s.d.=0.002) and Admixed American (ΔR^2^ = 0.065; s.d.=0.002) ancestry groups, predictive performance was similar to that of the AoU European group (**Fig. 4e**), demonstrating a certain degree of transferability for HLA_all. The SNP-based GRS was less predictive than the HLA-allele-based GRS in all ancestry groups, but was again similar between European subcohorts of UKB and AoU (**Extended Data Fig. 3**). These results provide evidence that a polygenic component underlies EBV viral load, which is largely driven by the MHC region and can be transferred across ancestries when calculated based on HLA-alleles.

The four GRS were then applied to the UKB disease target cohorts, whose participants had been excluded from the generation of GRS. Compared to the control cohort, highly significant associations were observed between an elevated HLA_MHC I GRS in patients with MS, and an elevated HLA_MHC II GRS in patients with RA (**Fig. 4f**). For MS, this effect was abolished when HLA-A*02:01, which is associated with MS^26^ and was among the most significant HLA-alleles in the GWAS, was excluded from the GRS (P(HLA_MHC I)=1.06×10^-4^, P(excl HLA-A*2:01)=0.056). In contrast, exclusion of HLA-DRB1*04:04 from the HLA_MHC II GRS did not abolish the association of this GRS with RA. We also observed a significantly lower HLA_all GRS in individuals with NHL (**Fig. 4f**).

Finally, we used the four GRS to conduct PheWAS in the European AoU-cohort, using 1,751 PheCodes (including the EBV-associated diseases listed above, except for Sjoegren disease; Methods; **Fig. 4g, Extended Data Fig. 3**, **Supplementary Table S20**). This PheWas replicated at P<0.001 all three significant associations identified in UKB, i.e., those of specific GRS with MS, NHL, and RA. However, in this hypothesis-free approach, the strongest associations were found for other disorders, namely T1D (beta = 0.176 for HLA_MHC II), inflammatory bowel disease (IBD, beta = -0.135 for HLA_all, and beta = -0.112 for HLA_MHC II), and hypothyroidism (beta = -0,039 for HLA_MHC I, beta = 0.037 for SNP_wo_MHC).

### Two-Sample Mendelian Randomization (2SMR)

To investigate whether EBVread+ (as exposure) has a causal effect on selected EBV-associated diseases, we performed 2SMR analyses (**Supplementary Table S21**, Methods). As outcomes, we used five diseases with strong evidence for epidemiological association (i.e., MS (case/ control and severity), RA, HL, NHL, SLE) and the three diseases identified by our PheWAS (i.e., IBD, T1D, hypothyroidism). We found suggestive evidence for causal effects of EBVread+ on RA (OR_wMed_ = 1.21, 95% CI = [1.09, 1.35]) and T1D (OR_wMed_ = 1.86, 95% CI = [1.64, 2.10]), which were consistent across multiple estimators (Methods, Supplementary Notes). However, significant heterogeneity of effects was observed across SNPs, and the causal effects for these outcomes were driven by SNPs in the MHC region (**Supplementary Table S22**, **Extended Data Fig. 4**, Supplementary Notes). No evidence for causal effects of EBVread+ were observed for any of the other seven outcomes (**Supplementary Fig. 9**), or for hair color, which was used as a negative control outcome.

## Discussion

Here, we established EBV-reads, obtained as a by-product of standard human GS, as a surrogate measure for EBV viral load in blood cells. While previous studies have demonstrated that viral reads of EBV are present in GS data^23^, and that GS data can be used to study disease associations of chromosomally integrated Human Herpes Virus 6^43^, our study is the first to demonstrate that GS-based EBV reads can be used as proxies for elevated EBV viral load, with high specificity.

Using this measure, we identified associations between EBVread+ and several non-genetic factors. Current smoking was identified as a modifiable factor that increases EBVread+ prevalence, consistent with data from oral samples^44^. Smoking is also a risk-factor for several EBV-associated diseases^45–47^, although the underlying mechanisms remain largely unknown. Research has shown that current smoking affects both adaptive and innate immunity, with the latter returning to normal upon smoking cessation^48^. We observed no effect of former smoking on increased EBVread+, suggesting a potential interaction of the innate immune system with current smoking status in EBV control. However, current smoking also increases memory B cell count^49^, which may also be an alternative explanation for the observed association.

At the genetic level, we generated evidence that EBVread+ is polygenic and characterized by a major (and largely equal) contribution of MHC class I and MHC class II. While associations at MHC class I and gene-set enrichment analyses implicated CD8+ cytotoxic T and NK cells, the association at MHC class II suggests that CD4+ T-helper cells play an important role in EBV control^42,50^. Genes implicated by common variants were enriched for IEIs, with a higher effect size for IEIs that lead to a loss of EBV-control (e.g., *CD70*)^51,52^. This implies that our analyses probably encompassed novel candidate genes for IEIs. One such candidate is *CD226*, which is a member of the immunoglobulin superfamily that contributes to NK and CD8+ T cell regulation^53^, and impairs CD8+ T cell response in chronic HIV when downregulated^54^.

Using genetically-predicted EBV viral load, we identified genetic overlap with two autoimmune-diseases with a previously reported association to EBV, i.e., MS and RA. While EBV is a prerequisite for MS, HLA-A*02:01, which reduces MS risk^26^, was among our most significant findings, and was associated with better EBV control. In contrast, no consistent effect on EBVread+ was found for the major MS risk allele HLA-DRB1*15:01^26^, suggesting a distinct biological mechanism. For RA, 2SMR also suggested a causal effect of EBV viral load. In RA cases, alleles at MHC class I were associated with lower, and those at MHC class II with higher, EBV viral load. This suggests a specific dysregulation of the immune response to EBV, rather than a generic loss of immune control of EBV. Our analyses also implicated EBV control in novel diseases, namely T1D, IBD / Ulcerative colitis, and hypothyroidism. For T1D, 2SMR suggested that EBV viral load might be a causal factor. The pathophysiological relevance of EBV may thus be broader than currently assumed.

Our study had several limitations. First, the detection limit imposed by standard GS coverage meant that the majority of individuals had a read count of zero. This prevented resolution of the lower end of the EBV viral load distribution, which would require specific quantitative measures (e.g., qPCR) or deeper GS data. We converted the zero-inflated EBV-read distribution into a binary measure, which enabled the use of standard statistical frameworks. Future analyses could apply quantitative approaches to increase statistical power, and individuals with extremely high read counts could inform future IEI studies. Second, despite controlling for confounders, unobserved factors may have confounded associations with EBVread+. However, this risk was reduced by stringent statistical adjustments and the use of data from two biobanks, which yielded largely similar results. Third, despite finding partial transferability of HLA-based GRS across ancestries, replication of the GWAS findings and downstream analyses in non-European ancestries is required. Finally, given the biological complexity of the MHC region, future studies must evaluate the functional mechanisms of genetic variants implicated in EBVread+ with respect to potential pleiotropic effects.

In summary, we have established EBV viral sequence traces from human GS data as the basis for future investigations into functional, mechanistic, and epidemiological aspects of EBV latent infection. Quantification of viral load based on host GS data can be applied to other human pathogens, opening new possibilities for the study of interactions between chronic infections and the host immune system in health and disease.

## Extended Data Figures

**Extended Data Figure 1:**
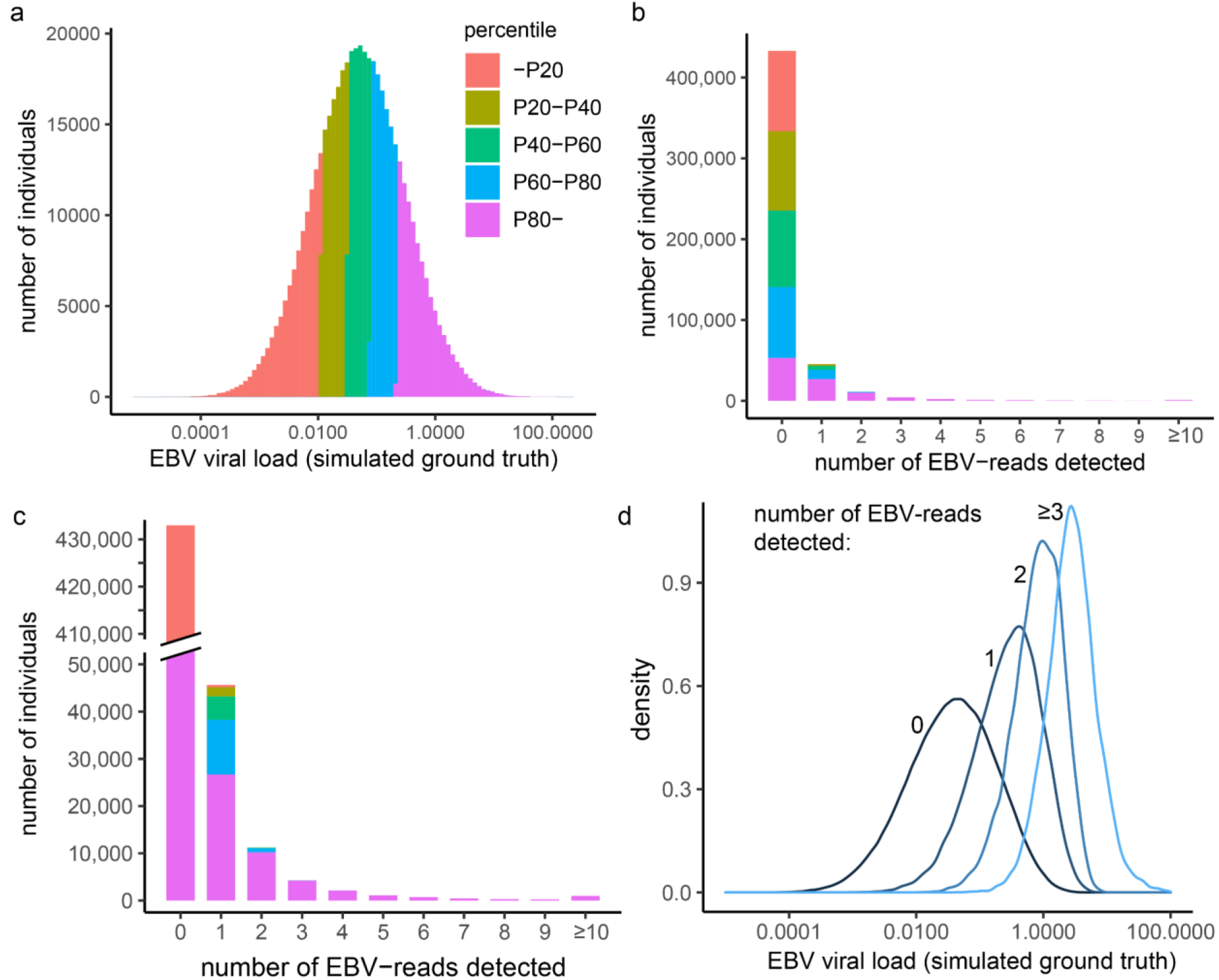
Simulation of EBV viral load and the generation of EBV-reads in GS. **a)** We modeled EBV viral load in 500,000 individuals using a log-normal distribution („ground truth“). This distribution was informed by prior observations on measured viral load in HIV^24^. The x-axis reflects theoretical units, which could be transferred to biological units if quantified standards were available. The numbers of individuals per unit are plotted on the y-axis. Individuals were assigned to 20% percentile groups (color coded). **b)** From the simulated viral loads, we then sampled “reads” for each individual, using a binomial distribution, with 400 million trials (approximately the average number of sequencing reads available per individual in our study). The probability values for successfully drawing EBV-reads were proportional to the viral load of the respective individual. Herefore, the success rate of the binomial distribution as well as the parameters of the log-normal distribution shown in **a)** were manually fitted to match the observed read count distribution in our data (Fig. 1c). **c)** is a zoom in on panel b. Note that the y-axis is interrupted for improved visualization. **d)** Within our simulation, EBV viral load increased with increasing numbers of observed EBV-reads (please note that reads of 3 or above are aggregated). Overall, this simulation demonstrates that a log-normal viral load distribution plausibly explains the observed distribution of the EBVread-counts in our data, and suggests that the number of detected EBV-reads is a measure of viral load (see also Supplementary Notes).

**Extended Data Figure 2:**
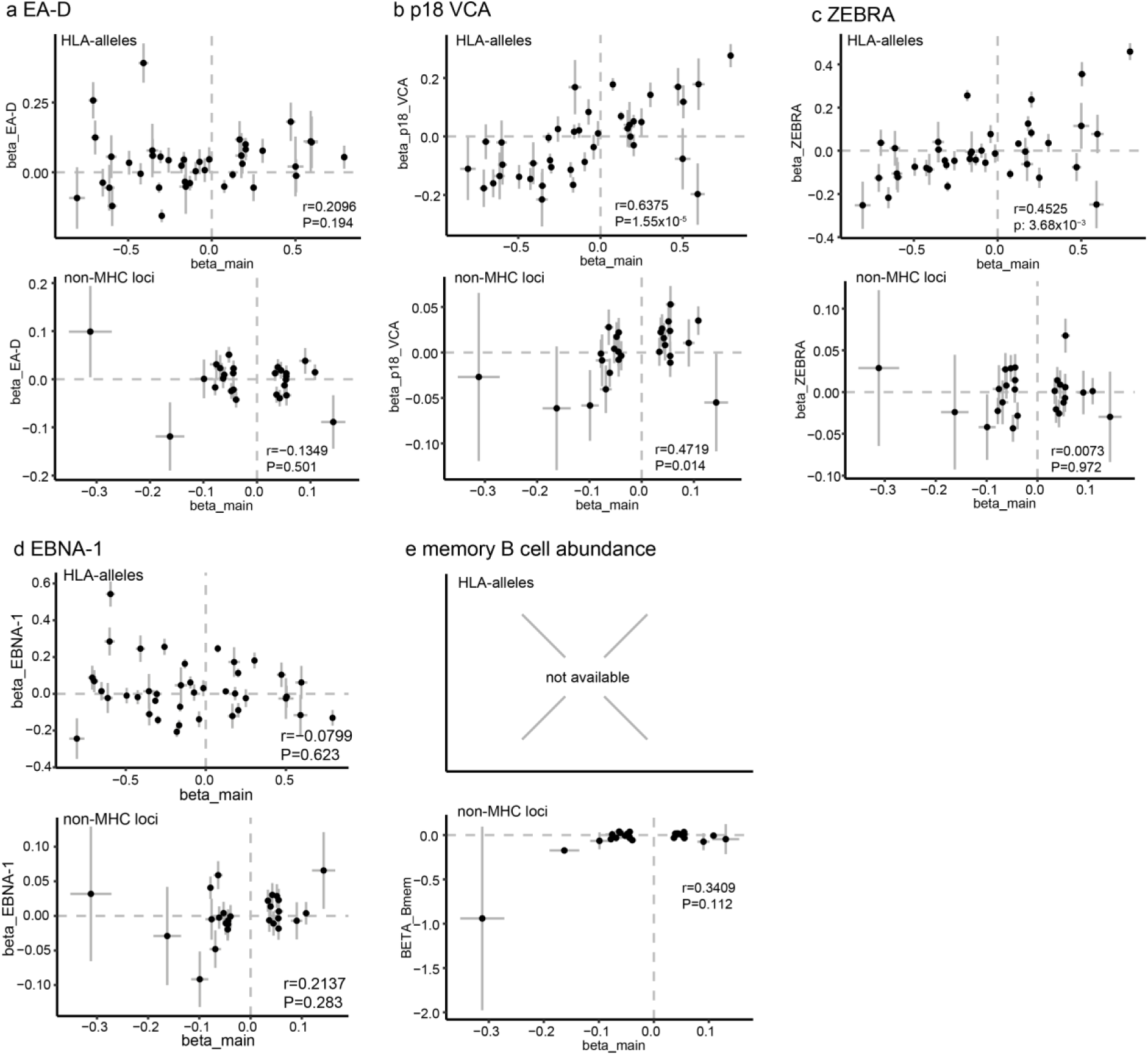
Comparison of effect sizes for lead variants at genome-wide significant loci of EBVread+ with selected traits. Effect sizes and alleles were retrieved from publicly available GWAS summary statistics of EBV antibody abundance (panels a-d)^29^ or abundance of memory B cells^30^ (panel e; no MHC data available) were plotted against betas of the EBVread+ association analysis. Only independent HLA-alleles with *P*<5×10^-8^ (upper panels) and lead variants at genome-wide significant non-MHC loci were used. To maximize the overlap (i.e., number of variants present in both summary statistics), SNPs in LD were chosen if lead SNPs were unavailable (see **Supplementary Tables S6, S9** for SNPs and HLA-alleles, respectively). Spearman correlation coefficients (r) and respective P-values (P) are indicated in the right-bottom corner of each panel.

**Extended Data Figure 3:**
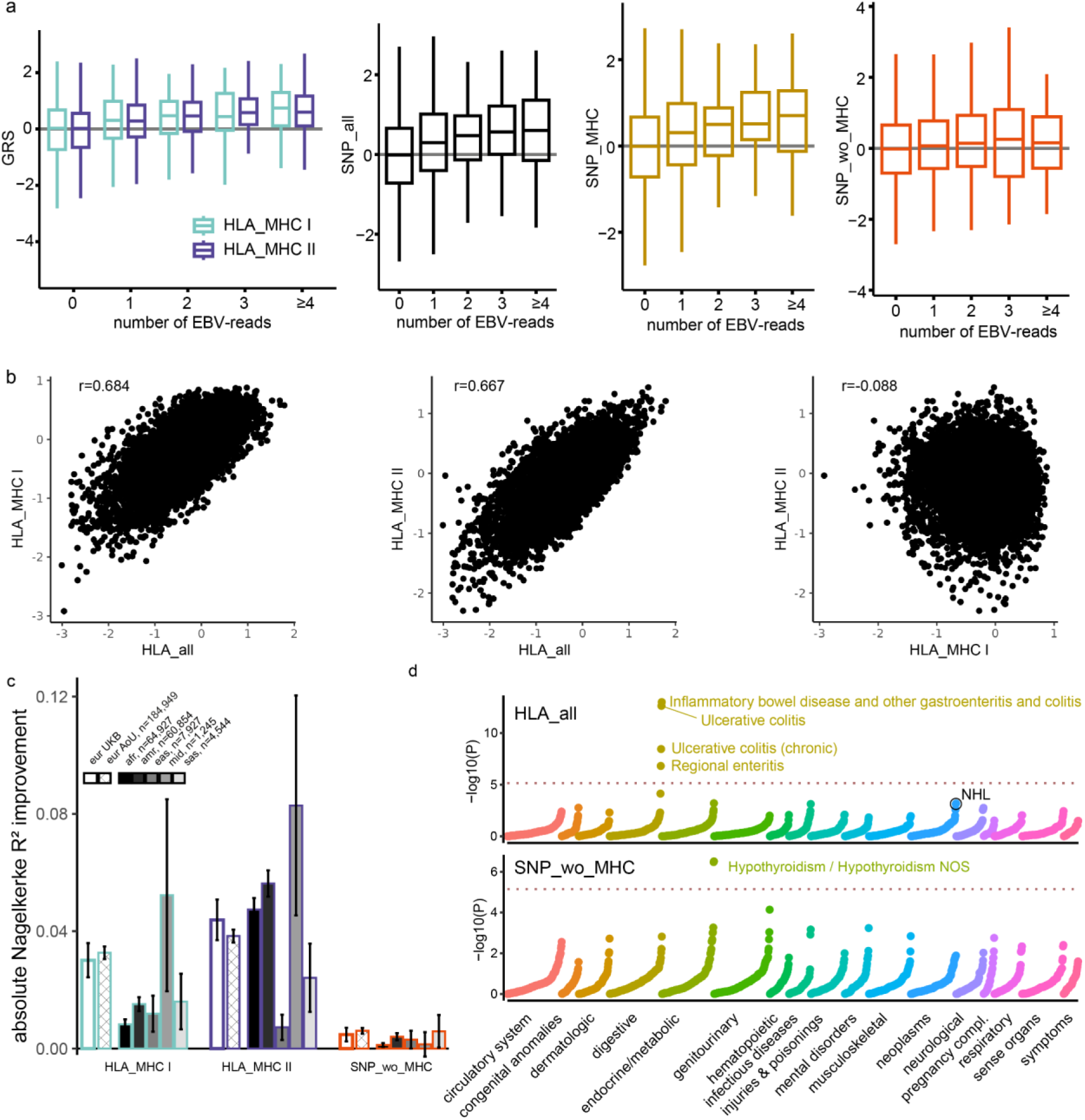
Prediction of EBVread+ using Genetic Risk Scores and PheWAS. **a)** Individuals from the serology control cohort (n=6,531) were stratified according to the number of EBV-reads detected in the GS data, and the distributions of specific GRSs within these groups were plotted. **b)** Scatter plots of individual GRSs (indicated by the axis labels) illustrate the correlation structures between HLA_all, HLA_MHC I and HLA_MHC II. Pearson correlation coefficients (r) are used to quantify correlations and indicate weak correlations between the GRS encompassing HLA-alleles from MHC class I vs those from MHC class II. **c)** In analogy to Fig. 4e, improvements in Nagelkerke’s R^2^ relative to base models within UK Biobank (taken leftmost bar of each GRS category), and six continental ancestries in AoU for the indicated GRSs are given. **d)** PheWAS using HLA_all and SNP_no_MHC in analogy to Fig. 4f. In addition to annotating all significant PheWAS associations, the association identified in UKB with NHL (HLA_all) is encircled.

**Extended Data Figure 4:**
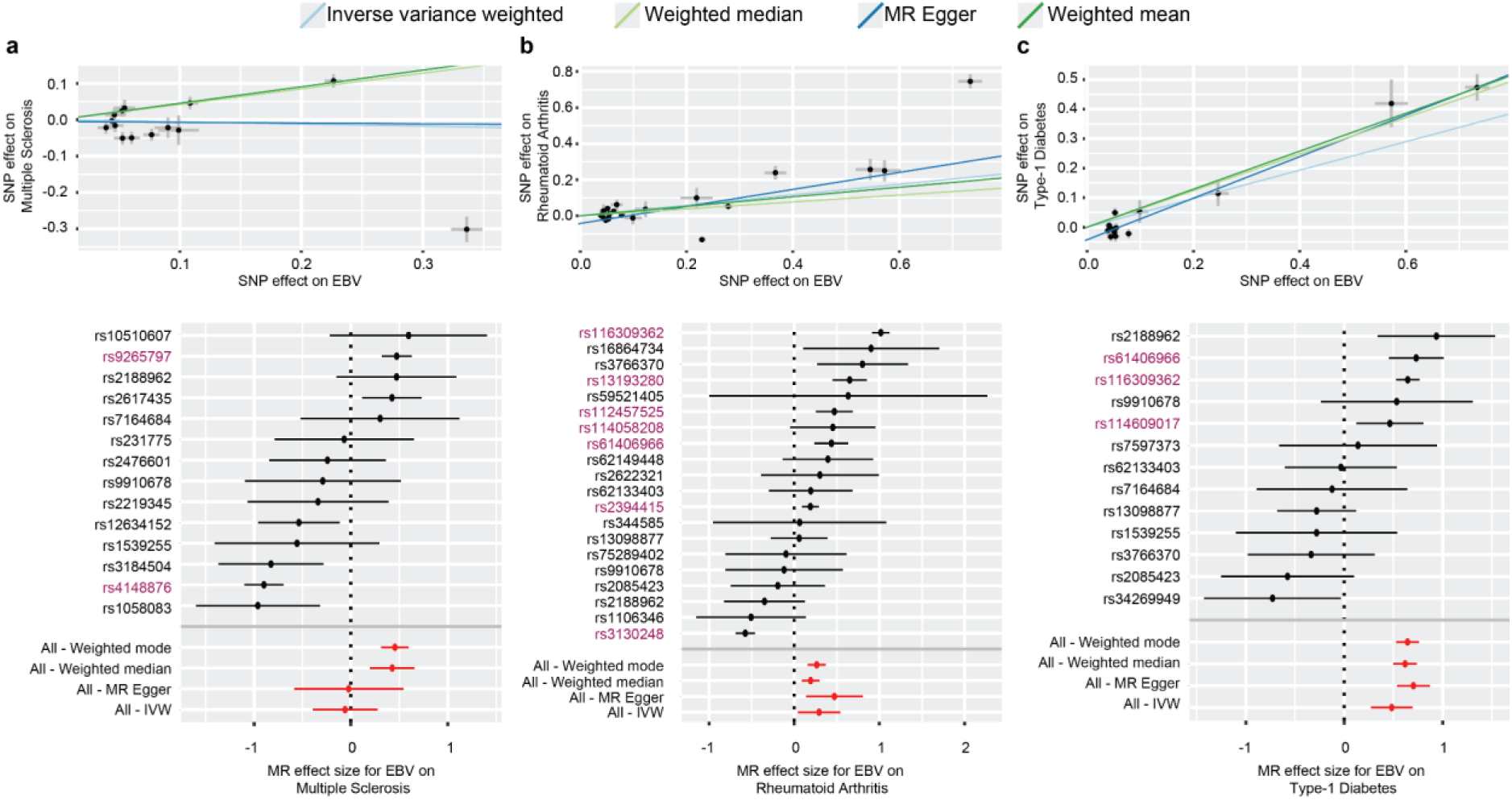
2SMR results for three traits that showed significant effects using two or more estimators. The figure shows scatter plots using the four standard estimators, as well as forest plots. MHC SNPs are highlighted in purple. **a)** Multiple sclerosis, **b)** Rheumatoid arthritis **c)** Type 1 diabetes.

## Methods

### Analysis of UK Biobank data

UK Biobank (UKB) Tier-3 data was accessed based on application-ID 135122. Analyses of individual level data were conducted within the UKB research analysis platform (RAP) using the frameworks R (v4.4.0), python (v3.9.16) and snakemake (v.32.4). Plots were generated using ggplot (v3.5.1 within tidyverse 2.0.0) unless otherwise indicated.

#### Identification of high-quality EBV-reads

All individuals of the UKB with available genome sequencing (GS) data were included in the initial stage of analysis. DNA extraction, library preparation, sequencing and alignment has been described elsewhere^55,56^. In short, DNA was extracted from the buffy coat of whole blood, PCR free libraries were generated using the NEBNext Ultra™ II PCR-free kit (NEB) and sequencing was performed with Illuminás NovaSeq6000 system on S4 flow cells (150 or 151 bp paired-end reads). Fastq data were then aligned using Illumina DRAGEN (v3.7.8) with a GRCh38 graph genome and provided as CRAM files (field 24048, described at medrxiv: https://www.medrxiv.org/content/10.1101/2023.12.06.23299426v1). EBV-reads were extracted from these CRAM files of n=490,293 individuals (*UKB*-cohort), using samtools (v1.20), and queried for reads that had been mapped to the EBV genome (NC_007605.1); specifically, to the contig chrEBV which was readily present within the GRCh38 reference genome. Only reads where both, forward and reverse, reads mapped to the EBV genome were retained. Reads were removed if they had more than 20 soft-clip bases, less than 120 bases matching the reference or were duplicate reads (see Supplementary Note 1 for the respective commands). These filter settings were determined by visual inspection of the aggregated alignment file of EBV-reads using the Integrative Genomics Viewer (IGV; v2.12.3), based on reads that showed evidence for unspecific mapping.

To rule out potential batch effects during library preparation, we calculated the proportion of individuals with EBV-reads per library prep plate (field 32056). We excluded plates with a high proportion of EBVread+ individuals (>2 standard deviations; nrm=3,979). Next, we excluded individuals with (i) low GS data quality as reported in data field 32064 (nrm=237), (ii) sex chromosome aneuploidies (field 22019) based on DNA array genotyping data, or (iii) discrepancies between reported and genetic sex (fields 31 (i) and 22001 (ii); nrm=5,553). Finally, for some analyses individuals were limited to those with European ancestry (nEUR=403,192, *EUR* cohort), selected based on UKB field 22006 (i.e. self-reported ’White British’ ethnicity) and very similar genetic ancestry based on a principal components analysis (PCA) of the genotypes. Individuals sero-positive for EBV were identified using the data field 23053.

#### Generation of HHV-7 sequencing read dataset

We generated a similar dataset for Human Herpesvirus 7 (HHV-7). Again, we extracted unmapped reads from the GS CRAM files and aligned them to an HHV-7 fasta file based on NCBI data (NC_001716.2), using bwa-mem2 (v2.2.1). Filters for sequencing reads were set to stricter criteria, due to a higher rate of unspecific mapping (*data not shown*). Specifically, we kept reads that met the following criteria: forward and reverse sequences mapping to HHV-7, non-zero mapping quality, 10 or less soft-clip bases and 130 or more reference bases, not marked as duplicate. Finally, only reads were counted whose forward and reverse read passed these filters (see Supplementary Note 1). Individuals sero-positive for HHV-7 were identified using the data field 23058.

#### Genetic variant sets for QC and association analyses

(a) For QC, we generated a high quality set of common variants for the *EUR*-cohort using plink v1.90b7.4^57^, by selecting genotyped variants (UKB field 22418) matching the following conditions: Variant minor allele frequency (MAF) above 1%, variant level missingness below 5%, Hardy-Weinberg p-value (midp-correction) above 1E-25, location on chromosome 1-22 and outside of long-range linkage disequilibrium (LD) regions (https://genome.sph.umich.edu/wiki/Regions_of_high_linkage_disequilibrium_(LD)). Finally, variants were pruned based on LD (plink parameter --indep-pairwise 1000 50 0.05), which resulted in a set of 80,217 variants. Within this variant set, individuals with a variant-missingness > 5% were removed (nrm=172 individuals). Related individuals (kinship-coefficients ≥ 0.0442) were identified according to the file “Genotype Results/Genotype calls/ukb_rel.dat” as provided by the UKB. To obtain an unrelated subcohort, related individuals were iteratively filtered out, starting with individuals with the highest numbers of relations (“non-rel.” subsets, see Fig. 1a). PCA was then conducted using FlashPCA (v2.0)^58^, on the unrelated subset of the cohort, while related individuals were subsequently projected onto the same principal component space. For regenie step 1 (see below), a slightly different variant set was obtained in analogy to the filter steps described for PCA, except that the MAF-threshold was set to 0.1% and the settings for pruning were relaxed (plink parameters: --indep-pairwise 1000 50 0.2). This resulted in 279,126 variants for 403,029 individuals. In total, 403,014 individuals had both principal components calculated and step1 variants available (*EUR*-cohort).
(b) For association analyses of common variants, we generated a set of variants that were previously imputed using the Haplotype Reference Consortium (HRC) and UK10K haplotype resource^20^ (UKB field 22828), with imputation info-scores above 0.8 (n=29,865,259 variants). Note that imputed data were missing for 481 individuals, which were not considered in the association analyses. For the HLA region we additionally accessed HLA-alleles which were imputed with the HLA*IMP:02 algorithm and provided within the UKB (field 22182). For compatibility with regenie step2, we converted the provided dosages to plink2 pgen-files.
(c) For rare-variant gene-based burden tests, we used exome-variants as provided by the UKB (field 23158). UKB exome sequencing and secondary analysis of sequencing data is described elsewhere^59^. In short, IDT’s xGen probe library was used for library preparation and sequencing was conducted as 75-base-pair paired end sequencing on Illumina NovaSeq 6000 platforms using S2 or S4 flow cells. The main steps of the secondary analysis comprised alignment to GRCh38 using bwa mem (v0.7.17), variant calling using deep variant (v0.10.0) and joint calling with GLnexus (v1.2.6). For quality control, only variant sites where 90 percent of individuals had a coverage of at least 10 were retained, as suggested.

#### Identification and preprocessing of covariates

We considered the following potential covariates that might confound associations with viral read counts: aspects of sequencing, technical biases, blood composition and demographic information (see **Supplementary Table S3**). While this selection is adopted from ref ^60^, neutrophil percentage is replaced by the strongly correlated lymphocyte percentage. Based on results from the initial phenome-wide screen for SNOMED-IDs associated with EBVread+ in the AoU cohort (see below), we additionally included smoking status, pack years of smoking, number of cigarettes smoked per day (or previously smoked in cigar/pipe smokers) and number of weekly alcoholic drinks. Covariates of the *EUR-cohort* were preprocessed to ensure suitability for association testing. First, individuals with missing values in blood count traits, in fasting time prior to blood sampling, smoking status and time of blood sampling for the initial visit of the assessment center (instance 0) were removed from further analysis (nrm=19,510). The number of cigarettes smoked per day (or previously smoked in cigar/pipe smokers), named CigDayCurrent, was extracted from the UKB fields 3456 and 6183, respectively. Missing values in current smokers were imputed with the mean of observed values (nimputed=10,726). The number of alcoholic drinks per week (DrnkWk) was calculated from weekly alcoholic drinks or, if not available, from monthly drinks or alcohol intake frequency, according to UKB. If all values were missing, mean imputation was performed (nimputed=233). Additionally, to ensure plausibility of covariates and in analogy to ref ^60^, outliers on four variables were winsorized at specific upper bounds: Fasting time value: 18; Pack years of smoking: 100; DrnkWk: 140; CigDayCurrent: 60. Next, individuals were excluded based on additional outlier measures: (i) individuals with very early or very late date of recruitment (outside of the range April 2007 to July 2010, nrm=849), (ii) individuals with outliers in blood count traits, i.e., any blood measurement with a Z-score > 4, based on reference^60^ (nrm=11,522).

Covariates that could not be reasonably approximated by a linear relationship with respect to the presence of EBV or HHV-7 reads were modeled with natural splines (R library splines, v4.4, see reference^60^. The natural splines were fit on the unrelated subset of individuals, for the following covariates and using the degrees of freedom (df) given in parentheses: date of attending assessment center (df=7); month of attending assessment center (n=3); time blood sample collected (df=5). With the fitted natural spline models we calculated the chance of observing EBV or HHV-7 reads for each individual and each of the aforementioned covariates, respectively. These transformed versions of the covariates were subsequently used instead of the raw values.

Finally, correlated covariates were identified by calculating Pearson correlations, where one of each pair of covariates with a correlation > 0.7 was removed, which led to the removal of four covariates. We calculated two additional covariates: age*sex and age*age, which resulted in 28 potential covariates (see **Supplementary Table S3**).

#### Selection and effects of covariates

To define the most relevant set of the 28 covariates, we used forward and backward selection with Bayesian information criterion (BIC) as a quality parameter (stepAIC function of the ‘MASS’ library, v7.3-61 within R). The phenotypes “EBVread+” and “HHV-7read+” were used as outcomes, respectively, and covariates were retained for the use in our genome-wide association studies (GWAS) if they were selected by one of the two analyses. The 18 covariates finally selected for GWAS are given in **Supplementary Table S3.** Further, the effects of changes in individual covariates were determined using marginal standardization^61^. To obtain covariate distributions, marginal standardization was applied to each of 1000 resamples (bootstrapping). Only non-related individuals of the respective cohorts were used.

#### Identification of immunosuppressive conditions and EBV-associated diseases

In order to reduce the effect of exogenous immunosuppression in our data, we identified individuals who had been taking immunosuppressive drugs (including individuals with prior transplants) or who were reported to be infected with HIV. For immunosuppressive drugs, we considered glucocorticoids and other immunosuppressants as given in **Supplementary Table S19**. Medication taken at the time of visit at the UKB assessment center (verbal interview, field 20003) was used to identify individuals taking the aforementioned drugs (nrm=9,681). Individuals with HIV infection were identified via UKB fields 130204,130206,130208,130210 and 130212 (nrm=230). For genetic risk analyses, additional EBV-associated diseases were identified on the basis of self-reporting by individuals in the assessment centre, International Statistical Classification of Diseases and Related Health Problems, 10th revision (ICD-10) codes, codes for operative procedures (OPCS4; **Supplementary Table S19**).

#### Association analysis of common variants and HLA-alleles

We conducted GWAS including related individuals, using regenie (v3.2.4)^62^. The 18 selected covariates and 20 PCs generated from common variants (see above, respectively) were used as covariates for viral read count related traits. For regenie step 1, the step1 variant set was used to account for relatedness. Regenie step 2 was applied to the step2 variant set and the following parameters were used in the main analysis: saddle point approximation (SPA) to account for case-control imbalance and a filter to only consider variants with a predicted minor allele count of 25 or higher. In the final EBVread+ summary statistics we observed that P-values for variants at the MHC region reached a plateau at -log10(P) of 306.653, likely reflecting statistical limitations of the SPA for extremely strong associations. We repeated the SNP-based association analysis for the MHC region (25Mb - 36Mb, chrom. 6) without SPA with the aim to get unbounded P-value estimates. Indeed, rerunning the GWAS without SPA for the MHC region revealed P-value estimates for all variants above the plateau, with all other P-values remaining strongly correlated and effect sizes remaining nearly identical for all variants with or without SPA (**Supplementary Fig. 10**). In addition, for association analysis of HLA-alleles, regenie step 1 and step 2 were conducted as described above, only the step 2 variant set was replaced by the imputed HLA-alleles.

#### Gene-based association analysis of rare variants

The gene-based association analysis of rare variants (RVASgene) was performed in analogy to the association analysis of common variants with identical phenotype and covariate files, and the same procedure for regenie step 1. However, for regenie step 2, we used SKAT-O as a test (parameter: ‘--vc-tests skato’) and restricted the analysis to rare variants with an alternative allele frequency below 1% (parameter ’--vc-maxAAF 0.01’). Very rare variants (minor allele count < 10) were collapsed into burden masks using the parameter ’--vc-MACthr 10’. Firth correction was applied to effect sizes, with parameters ’--firth’, ’--firth-se’, and ’--approx’.

Variant annotations and assignment to genes were used as provided within the UKB RAP (file: ukb23158_500k_OQFE.annotations.txt.gz), based on ref ^59^. In short, the annotation was created using SnpEff with Ensembl v85 as the underlying database. Missense variants were classified based on the number of missense prediction scores indicating a deleterious effect: 5/5 – ‘likely deleterious’, 1–4/5 – ‘possibly deleterious’, and 0/5 – ‘likely benign’. The scores used were SIFT, PolyPhen2 HDIV, PolyPhen2 HVAR, LRT, and MutationTaster.

The following definitions of variant pathogenicity (masks) were used within the gene-based association analysis of rare variants: 1) M1: predicted loss-of-function variants, 2) strong_coding: variants from 1) and likely deleterious missense variants, 3) medium_coding: variants from 2) and possibly deleterious missense variants, and 4) all_coding variants from 3) and likely benign missense variants.

### Analysis in the All of Us cohort

We used release 8 (C2024Q3R3) of the All of Us (AoU) cohort, which included array and GS data generated from DNA sampled from blood of 365,931 individuals (*AoU*-cohort). For a thorough description of the AoU resource, including data generation, processing and quality control of genomic data, please refer to reference ^21^ and accompanying documents.

#### Generation of EBV-read data and cohort from GS data

First, EBV-reads were extracted from CRAM-files of participants of the AoU program as described for UKB participants. At individual level, we restricted our analyses to individuals who were unrelated, without mismatch between reported and genetic sex and who were not flagged as population outliers (“flagged samples”) in accompanying documents (*AoU-QC-*cohort, n=336,123). In addition, we only included individuals with plausible time points of DNA sampling. For population-specific analyses the precomputed genetically predicted population backgrounds were used, which assigned each individual to one of six continental populations (African, Admixed American, East Asian, European, Middle Eastern, South Asian; **Supplementary Fig. 2**; see Genomic Research Data Quality Report).

#### Phenome-wide screen for SNOMED-IDs associated with EBVread+

We used the *AoU-QC*-cohort and additionally filtered for individuals with electronic health record (EHR) data available and who did not have extreme times of sample collection (within the time-frame 11:00 AM to 11:59 PM; n=266,186). We only considered Systematized Nomenclature of Human and Veterinary Medicine (SNOMED) concept IDs that appeared in at least 250 individuals within the health event dataset (resulting in 11,111 SNOMED concept IDs). These were tested for associations with the presence of EBV-reads. We used logistic regression models with the presence of EBV-reads as outcome, the presence of a SNOMED ID as predictor, respectively, and age, sex, age*sex and 16 PCs capturing population background as covariates based on information provided by AoU in their helper files accompanying GS data. In a second step, we also included HIV- and smoking-status as covariates. Likelihood-ratio test was used to calculate p-values on models with or without the respective SNOMED concept ID as predictor.

#### Variant sets and individuals for PCA and genetic analyses

Array data contained 1,739,269 variant sites and were accessed as plink 1 files. Short-read GS data were accessed as plink2 binary files, multiallelic sites were split into biallelic records using bcftools norm (v1.12). Variant level missingness and Hardy Weinberg equilibrium statistics were calculated using plink (v1.90b6.22). While information on HLA-alleles were readily available in UKB, this is not the case in AoU. Therefore, alleles of HLA genes were imputed using the HLA-TAPAS framework^63^. We created a reference panel for the AoU cohort using the MakeReference module of HLA-TAPAS with genotype data of the 1000 genomes project phase 3 (1000Gv3; n=2,504 individuals) and accompanying HLA-alleles (provided by HLA-TAPAS). HLA-alleles were imputed with the SNP2HLA module of HLA-TAPAS using the genotyping array data of AoU and batches of 2000 individuals.

The identification and analyses of principal components was similar to the UKB procedure, except that PCs with 20 dimensions were calculated within individuals of each of the six continental populations. For further analyses we selected individuals without reported HIV infection. Individuals with outlying timepoints of blood draw were excluded (blood draw time before 11 am).

#### Genetic association studies

Within each population subcohort, association analysis was performed using regenie (v2.0.2) without using step 1. All other command line parameters given to regenie were identical to the analysis conducted within UKB. We selected similar covariates as in the analysis within UKB, i.e.: sex, age, age*sex, mean sequencing coverage, probability of EBVread+ predicted based on the hour as well as the week of biosample collection and (natural spline with 5 and 6 degrees of freedom, respectively), nicotine usage, sequencing site and 20 principal components. Please note that certain covariates, such as blood count traits, are not directly available in AoU and therefore could not be included. Here, we report the results for the European population.

#### Definition of smoking- and HIV-status

Current smokers were defined as individuals that gave the answer “Some Days” or “Every Day” to the question “Do you now smoke cigarettes every day, some days, or not at all?” of the AoU Lifestyle questionnaire or as the presence of “Nicotine dependence” within the conditions. Individuals were annotated as having HIV if any of the following was true: Answered “Yes” to the question “Are you still seeing a doctor or health care provider for HIV/AIDS?”, gave any valid age range as answer to the question “About how old were you when you were first told you had HIV/AIDS?” or was annotated with any of the following conditions: Human immunodeficiency virus infection, Human immunodeficiency virus carrier, HIV-positive, Acute HIV infection, AIDS or AIDS-associated disorder.

### Genetic risk loci associated with EBVread+

#### Annotation of non-MHC risk loci

Summary statistics were uploaded into private instances of LocusZoom^64^, which was also used for generation of Regional Association Plots^64^ and FUMA. Genome-wide significance was defined as P<5×10^-08^, and independent risk loci were defined based on FUMA’s SNP2GENE module (v1.6.3)^39^, which groups variants into genomic risk loci based on LD structure (1000Gv3, European population), using an R² threshold of 0.6 without MAF cutoff and a lead SNPs merging distance of 250 kb. For annotation of loci we used either the statistical lead variant (**Supplementary Table S4**) or a proxy-variant as listed in **Supplementary Table S9**. For each locus we report (i) closest gene (based on distance of lead SNP to the transcription starting site), (ii) LD genes (based on positional location in regions harbouring variants with R²>0.2 to lead SNP), (iii) eGenes from GTEx (based on GTAdult GTEx v10, with genome-wide significant single-tissue eQTL effects (P<5×10^-8^) in any tissue), and (iv) V2G scores from Open Targets v22.10 (based on a cutoff >0.1). We considered evidence from at least three out of four approaches as consistent evidence. To identify pleiotropic effects of lead variants **Supplementary Table S9**, we used OpenTargets v22.10 to retrieve all traits at P<0.005 that were reported in either GWAS Catalog, UKB or Finngen. To identify potential targets for drug repurposing, approved drugs (clinical phase IV) targeting the identified genes were retrieved from OpenTargets v25.3.

#### Generation of credible SNP sets

Fine-mapping for each non-MHC locus was performed with SuSie ("Sum of Single Effects Regression")^65^, which uses Bayesian statistics to determine the posterior inclusion probability (PIP) for each SNP within a credible set (CS, defined as the region harbouring cumulative PIPs >0.95). Fine-mapped regions included 1 Mb as standard, based on haplotype blocks and recombination rates at individual loci. For two loci, 12q24.12_SH2B3 and 5q31.1_SLC22A5, regions were extended due to particularly long extents of associated haplotype blocks, to 3 Mb and 5 Mb, respectively. The LD matrix required for PIP determination or colocalization analysis was generated from the imputed genotype data of an unrelated subset of European UKB individuals (n=339,539; filtered as described for the *EUR*-cohort; however, the filter for available principal components was not applied) with plink2 (v2.0.0-a.6). All variants within the credible SNP sets were annotated using Ensembl Variant Effect Predictor^66^ (VEP; release 113) for GRCh37 to identify coding variants. For those, information was also retrieved from the ClinVar database (version June, 2023)^67^ and AlphaMissense prediction scores^68^ are provided.

#### Association of loci in HHV-7 and B-memory cells

Lead variants of loci associated with EBVread positivity were analyzed in HHV-7 association data (generated as described above) and in a previously published GWAS on memory B cells^30^ (GCST90001407). For HHV-7, to distinguish loci with shared causal variants from those where distinct variants underlie the observed association, colocalization analysis was performed at non-MHC loci showing nominal significance using coloc (v5.2.3)^69^ in R (v4.4.2), and H3 and H4 probabilities are reported.

#### Identification of single-cell eQTLs

To investigate potential regulatory effects at the 27 non-MHC loci, lead SNPs based on **Supplementary Table S9** were looked up in the PBMC single-cell RNA sequencing dataset from “OneK1K” cohort^32^. Specifically, we retrieved all eQTLs reported for SNPs located within each genomic locus, and computed LD with the corresponding GWAS lead SNPs using an LD matrix derived from the European population (based on LDmatrix^70^, 1000Gv3, EUR). eQTL effects were reported for SNPs with R^2^ > 0.7.

### Gene-level annotations and enrichment analyses

*Gene-based association analyses of common variants.* MAGMA v1.08^36^ as implemented in FUMA^39^ was used to jointly test associations between SNPs and genes they were annotated with, generating gene-level association statistics. All analyses in FUMA were performed using default FUMA settings, except for the modifications specified below. First, SNPs were annotated to genes using the MAGMA gene boundaries Ensembl v102 file (excluding the extended MHC region as previously suggested^60^ (25-36 Mb), to define gene locations and the European 1000Gv3 genotype data and to provide LD information for SNP-to-gene mapping and gene-level association testing. A window of 10 kb upstream and 1.5 kb downstream was used for assigning SNPs to genes. Gene-level testing was performed based on the GWAS summary statistics of EBVread+, and association P-values were corrected for the number of tested genes (n=19,736). Genes with Bonferroni-corrected P-values<0.05 were considered significantly associated with EBVread+. We used two target gene sets defined by genes underlying inborn errors of immunity (IEI) to test for their enrichment among the genes associated with common variants for EBVread+. Specifically, we used 14 genes underlying monogenic forms of EBV-susceptibility^15^ and 483 genes listed as IEIs (based IEI classification October 2024, document “IUIS-IEI-listJuly2024v2.xls”, downloaded from https://iuis.org/committees/iei/ on May 6th, 2025), of which n=456 were available in our data. The full MAGMA gene list was additionally used as input for the following enrichment analyses as described below. Results were visualized using R.

#### Enrichment analyses

MAGMA as implemented in FUMA’s SNP2GENE module^39^ was used to relate genes associated with EBVread+ (i) to Gene Ontology Biological Processes (GOBP) via gene-set analysis and (ii) to different tissues via gene-property analysis of gene expression. The gene-set analysis included 7,743 GOBP terms; the gene-property analysis included 54 distinct tissue types from GTEx v8. For each of the two approaches, significance thresholds were adjusted by Bonferroni-correction to control the family-wise error < 0.05.

#### scDRS analysis

Single-cell disease relevance score (scDRS) analyses were performed using scDRS v1.0.3^41^. scDRS identifies cell types within a single-cell RNA sequencing (RNAseq) dataset via excess expression of genes from the EBVread+ GWAS that are prioritized by MAGMA (see above). We downloaded single-cell RNAseq data from peripheral mononuclear blood cells (PBMCs) from the 1M-scBloodNL project, published by the sc-eQTLGen consortium^40^. QC-ed raw gene expression counts of the samples processed with 10x Genomics v3 as well as the corresponding genes, barcodes, and cell type annotations were downloaded on January 30, 2025. The dataset contained two levels of cell type annotations (level 1: 10 cell types; level 2: 26 cell types), of which we focused on the broader annotation level for the main analyses. Using the Seurat package v5.2.1 in R v4.3.2, the scRNAseq dataset was filtered for the subset of untreated cells and for level 1 cell type clusters containing more than 100 cells, leading to the exclusion of cells annotated as “unknown” (77 cells) or “plasma B” (80 cells). 37,033 cells annotated to 8 cell types remained for the scDRS analysis. The dataset was written to a H5AD file for input to scDRS using the SeuratDisk package v0.0.0.9021. The top 1000 EBVread+ MAGMA genes and their z-scores as weights were obtained using the scdrs munge-gs command. The scdrs compute_score command was used with default parameters on the prepared scRNAseq dataset and MAGMA gene set to calculate scDRS. Group analyses (cell type association and heterogeneity) for both cell type level 1 and 2 were conducted with the scDRS perform_downstream command, with default parameters. Multiple testing correction of *p*-values for the number of cell types within the respective annotation level was performed using the Benjamini-Hochberg procedure for controlling the false discovery rate.

### Genetic risk score analyses in UKB and AoU

#### Polygenic contribution to serology and EBV-associated diseases in UKB

To study the joint contribution of common variants associated with EBVread positivity to EBV-associated diseases within UKB, we first removed from the *No-outlier*-cohort cohort those individuals that were either included in the serology cohort, or annotated with EBV-associated diseases (see above and **Supplementary Table S19**). This cohort was used as “base cohort” for calculation of the genetic risk scores (GRS), while removed individuals were grouped into different target-disease cohorts (see **Fig. 1a**). To ensure independence of target and base dataset and to ensure independent observations within the target cohort, we removed all individuals that were related to any other individual of the target or the base cohort.

To generate SNP-based GRS, first an association analysis of common variants was performed within the base cohort as described above, except that only autosomal and genotyped variants with a minor allele count above 25 were used for regenie step 2 (n=739,066; for the sake of computational efficiency). Polygenic risk scoring was then performed using PRS-CS (v1.0.0)^71^, which included filtering the summary statistics for non-ambiguous SNPs. To generate separate GRS for the MHC regions and the non-MHC regions, the summary statistics generated using the base-dataset were additionally filtered to only contain the required regions (i.e. MHC region, including separate subsets of MHC classes I and II; non-MHC regions). Scores of individuals of the target-cohort were obtained using the score function of plink (v1.90b6.21).

GRS based on imputed HLA-alleles were calculated by fitting a multivariable logistic regression model to the base-dataset, where EBVread+ was the outcome. As predictors we used the 18 covariates and the 20 principal components that were also used in the associated analysis of common variants above, as well as 178 HLA-alleles that had a minor allele frequency of greater than 0.1%. After model fit, the coefficient estimates of the 178 HLA-alleles were retrieved. Scores for individuals in the target-cohort were generated by multiplying imputed HLA-allele dosages with coefficient estimates and summing these values. To obtain MHC class 1 or MHC class 2 specific scores, we calculated the GRS using the same coefficient estimates, but considered only the HLA-alleles belonging to the respective MHC class.

All risk scores were normalized to means of 0 and standard deviations of 1 within the combined target cohorts, to improve comparability.

#### Evaluation of GRS performance

We evaluated the performance of different GRS using individuals with serology data. Specifically, we fitted logistic regression models with the outcome EBVread+ and covariates only (age, sex, age*sex, 20 PCs; base model) or covariates and one of the GRS as predictors. We subsequently calculated Nagelkerke R^2^ for the base model and for models including GRS. To evaluate the variability of Nagelkerke R^2^ estimates, we used bootstrapping (n=1000). To test for significant differences between GRS values of two groups of individuals (e.g. the serology cohort and a disease cohort), we used the likelihood ratio test. As input for this statistical test, we fitted two logistic regression models to the individual level data of both groups and used an indicator variable of the group membership as outcome. The first model used covariates only as predictors (see above) and the second model used covariates and the GRS as predictors.

#### Transferability of GRS across populations and cohorts

We calculated SNP-based and HLA-allele-based GRS within AoU to check the transferability of our UKB-based GRS. For SNP-based GRS we used 1,509,024 genotyped variants within AoU, which had a HWE P-value ≥ 1×10^-10^ and a variant level missingness < 0.05 in each continental ancestry. We then used our EBVread+ summary statistics from UKB, which was calculated on imputed genotypes, as the base dataset. Polygenic risk scoring was then performed using PRS-CS as described above. To obtain HLA-based genetic risk scores, we used coefficient estimates of the 178 HLA-alleles from UKB (see above) and multiplied them with the estimated dosage for each HLA-allele in AoU. The mapping of HLA-allele names between the names given by HLA*IMP:02 and HLA-TAPAS was performed manually, which resulted in a successful mapping of 166 of 178 alleles (no clear mapping for one MHC class I allele and 11 MHC class II alleles).

#### Phenome-wide association of EBV GRS with PheCodes

We used the software package PheTK (v0.1.47)^72^ to assign PheCodes (v1.2) to individuals of AoU. Individuals were considered as having a certain PheCode if the PheCode was annotated at least twice to the individual, as suggested by PheTK. We used logistic regression models with the presence of a PheCode as outcome and GRS as predictors, respectively. Age, sex, age*sex and 20 population specific PCs were used as covariates. P-values were calculated using the likelihood-ratio test comparing models with and without the respective GRS. To comply with AoU publishing guidelines, we only report PheCodes annotated to >20 individuals, do not supply count related data and only give proportions when all underlying groups contain > 20 individuals. This resulted in an analysis of 1,751 PheCodes.

### Two-Sample Mendelian Randomization analysis

*Selection of outcome traits.* To test potential causal effects of genetically-predicted high EBV viral load on EBV-associated diseases, we performed Two-Sample Mendelian Randomization (2SMR) in R (v4.5.0) using the R packages *ieugwasr* (v1.0.3) and *TwoSampleMR* (v0.6.15)^73,74^. We examined causal effects between EBVread+ (exposure) and the following diseases (outcomes), based on different levels of evidence and using publicly available GWAS summary statistics: *(i) known EBV-associated diseases*: multiple sclerosis (MS) case-control^75^, MS severity^76^, Hodgkin’s disease^77^, non-Hodgkin lymphoma^77^, SLE^78^ and RA^79^. *(ii) candidate diseases based on significant PheWAS results:* hypothyroidism^77^, type 1 diabetes^80^, inflammatory bowel disease^81^. For each of the nine outcomes, GWAS selected did not include UKB samples and contained individuals of European ancestry, and summary statistics were retrieved from the GWAS catalog except for MS severity, where summary statistics were shared by the authors. Further details on the outcome GWAS are provided in **Supplementary Table S21**.

*2SMR.* We applied consistent QC on all GWAS summary statistics (exposure and outcomes), removing duplicate SNPs and retaining autosomal SNPs with MAF >0.01 and INFO score >0.80, if available. Genome-wide significant SNPs from our EBVread+ GWAS (exposure) were further filtered using LD clumping as incorporated in the ld clump() function with standard parameters, followed by harmonization with the outcome GWAS summary statistics (aligning strand, removing unresolvable palindromic SNPs). After variant filtering, LD clumping and harmonization, using genome-wide significant and independent loci of our EBVread+ GWAS summary statistics (UKB) did not indicate weak-instrument bias (*I*^2^ statistic = 0.99). We further used Steiger filtering^73^ to exclude potentially invalid instruments (SNPs showing stronger associations with the outcome vs with the exposure). 2SMR was performed using four different estimators: the inverse variance weighted (IVW) estimator, MR-Egger, weighted median and weighted mode. We used additional sensitivity analyses to describe the observed MR patterns, such as heterogeneity (Cochran’s Q statistic^82^) and pleiotropy tests (e.g., MR-Egger intercept^83^) and leave-one-out analyses.

Exposure-outcome associations that suggested a significant causal effect (P < 0.05/9) in ≥2 estimators were followed up using the more robust methods MR Robust Adjusted Profile Score (MR-RAPS, 10.1214/19-AOS1866) and MR pleiotropy residual sum and outlier test (MR-PRESSO^84^). Associations that showed (i) significant effects in < 3/6 estimators, or (ii) inconsistent effect directions were interpreted as not-likely to be causal. For associations with evidence for causality, we repeated the analyses after excluding SNPs in the MHC region. In addition, we used “red hair colour”^85^ (including UKB) as negative control outcome.

## Acknowledgements

We thank Anna Vyvers and Alexander Dilthey for critical discussions, and Christine Schmäl for manuscript editing. UK Biobank analyses were performed under application 135122. This work uses data provided by patients and collected by the NHS as part of their care and support, and we thank the participants and coordinators of the UK Biobank study. We also gratefully acknowledge All of Us participants for their contributions, without whom this research would not have been possible. We also thank the National Institutes of Health’s All of Us Research Program for making available the participant data examined in this study. We thank the International Multiple Sclerosis Genetics Consortium (IMSGC) for providing summary statistics on multiple sclerosis.

## Author Contributions

Conceptualization: AS, MMN, KUL

Methodology: AS, TMA, LF, SKH, FSD

Formal analysis: AS, TMA, LF, SR, MS, FSD

Data curation: AS, TMA, SR, ECB

Writing - Original Draft: AS, LF, ECB, KUL

Writing - Review & Editing: TMA, SR, MS, FSD, KB, AKP,MMN

Visualization: AS, TMA, LF, SR, MS, FSD, ECB

Supervision: MMN, KUL

Project administration: AS, KUL

Funding aquisition: KUL

## Ethics declaration

This study used de-identified data available from UK Biobank (UKB) and All of Us (AoU) which were accessed through their respective computing platforms. UKB has approval from the North West Multi-centre Research Ethics Committee (MREC) as a Research Tissue Bank (RTB). This approval allows approved researchers to operate under the RTB approval without requiring separate ethical clearance. The data collection of the AoU Research Program was conducted under centralized approval by the Institutional Review Board (IRB) of the AoU Research Program, with informed consent obtained from participants. This study adhered to the ethical guidelines outlined in the All of Us Data User Code of Conduct.

## Data availability

All genetic and phenotype data are available upon application and approved data access from the UK Biobank study and AllofUs project. All researchers will be able to access the data in the same manner that the authors did, including usage of the UKB research analysis platform and AoU workbench environments for the analysis of de-identified individual-level data. Complementary data used for secondary analyses were obtained from: OneK1K (https://onek1k.org/), eQTLgen 1M-scBloodNL (https://www.eqtlgen.org/sc/datasets/1m-scbloodnl-dataset.html), GTEx (https://www.gtexportal.org/home/), OpenTargets (https://platform.opentargets.org/), IUIS (https://iuis.org/committees/iei/), GWAS Catalogue (https://www.ebi.ac.uk/gwas/), the International Multiple Sclerosis Genomics consortium (https://imsgc.net/). All other data produced in the present study are available upon reasonable request to the authors.

## Code availability

Code used to extract EBV or HHV-7 reads from UKB and AoU is provided as Suppl Note 1. For all other analyses, publicly available software and tools were used which can be accessed as follows and which are referenced within the respective Methods section.

R v4.4.0 (https://cran.r-project.org/)

python v3.9.16 (https://www.python.org/)

snakemake v7.32.4 (https://snakemake.github.io/)

ggplot v3.5.1 (https://github.com/tidyverse/ggplot2/releases)

plink v1.90b7.4 (https://www.cog-genomics.org/plink/)

FlashPCA v2.0 (https://github.com/gabraham/flashpca)

regenie v2.0.2 / v3.2.4 (https://github.com/rgcgithub/regenie)

HLA-TAPAS (https://github.com/immunogenomics/HLA-TAPAS)

LocusZoom (http://locuszoom.sph.umich.edu//)

FUMA SNP2GENE v1.6.3 (https://fuma.ctglab.nl/)

MAGMA v1.08 (https://ctg.cncr.nl/software/magma)

SuSie (https://github.com/stephenslab/susieR)

VEP, Esembl release 113 (https://www.ensembl.org/info/docs/tools/vep/script/index.html)

coloc v5.2.3 (https://github.com/cran/coloc)

scDRS v1.0.3 (https://github.com/martinjzhang/scDRS)

PheTK v0.1.47 (https://github.com/nhgritctran/PheTK)

ieugwasr v1.0.3 (https://github.com/MRCIEU/ieugwasr)

TwoSampleMR v0.6.15 (https://github.com/MRCIEU/TwoSampleMR)

MR-RAPS (https://github.com/qingyuanzhao/mr.raps)

MR-PRESSO (https://github.com/rondolab/MR-PRESSO

LD score regression: https://github.com/bulik/ldsc

