## Supplementary Figures for "Host control of latent Epstein-Barr virus infection"

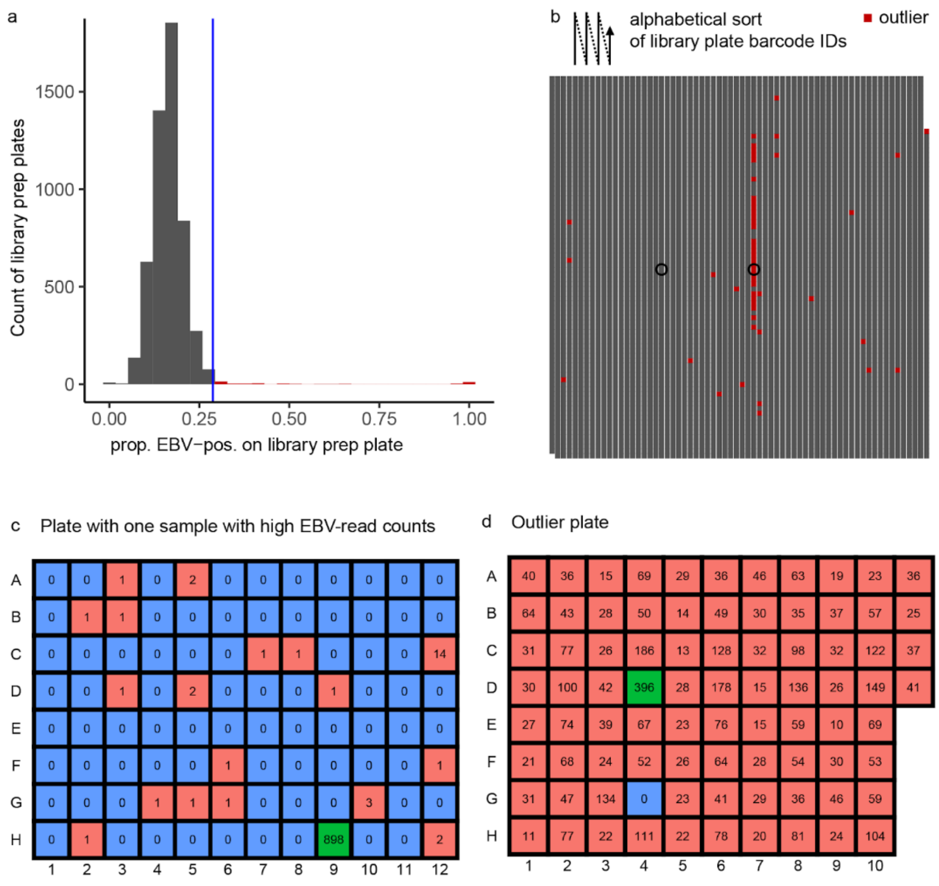


**Supplementary Figure 1: Identification of library plate outliers in UK Biobank. a)** The distribution of EBVread+ individuals per 96-well plate (library) was used to identify 51 library plates with high rates of EBVread+ individuals (above 28.8 % of samples with ≥1 reads, indicated by the blue vertical line). **b)** When library plate barcode IDs were sorted alphabetically, we observed a cluster of plates with high rates of EBVread+ (highlighted in red), probably illustrating batch effects. The two library plates highlighted by circles are shown exemplarily in **c** (one sample with high read count) and **d** (plate with high number of EBVread+ individuals. **c)+d)** Plate layouts with EBV-read counts for each sample are shown, with positions without EBV-reads being colored in blue while those with at least one EBV-read are colored in red. At each plate, the position with the highest EBV-read count is highlighted in green.

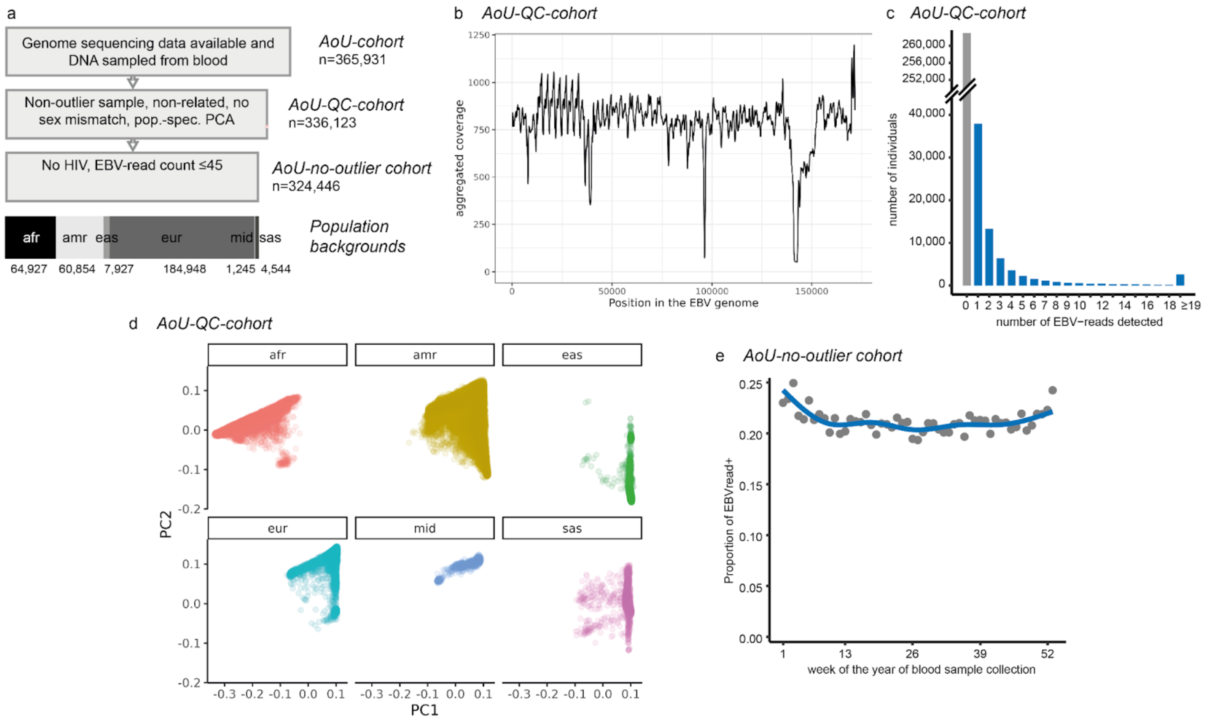


**Supplementary Figure 2: Analysis of EBVread positivity in All of Us cohort. a)** Flow diagram showing the generation of different All of Us (AoU) cohorts. Details can be found in the Methods section. The number of individuals in each of six population backgrounds for the AoU *no-outlier* cohort are given below the FlowChart, split by shades of grey. **b)** and **c)** are in analogy to Figures 1b and 1c of the main text. **d)** The first two principal components (PCs) of common genotypes as provided by AoU are displayed for each of the six population backgrounds. **e)** EBVread+ in relation to the week of the year in which blood samples were collected displays a seasonal pattern. Abbreviations: afr: African, amr: Admixed American, eas: East Asian, eur: European, mid: Middle Eastern, sas: South Asian.

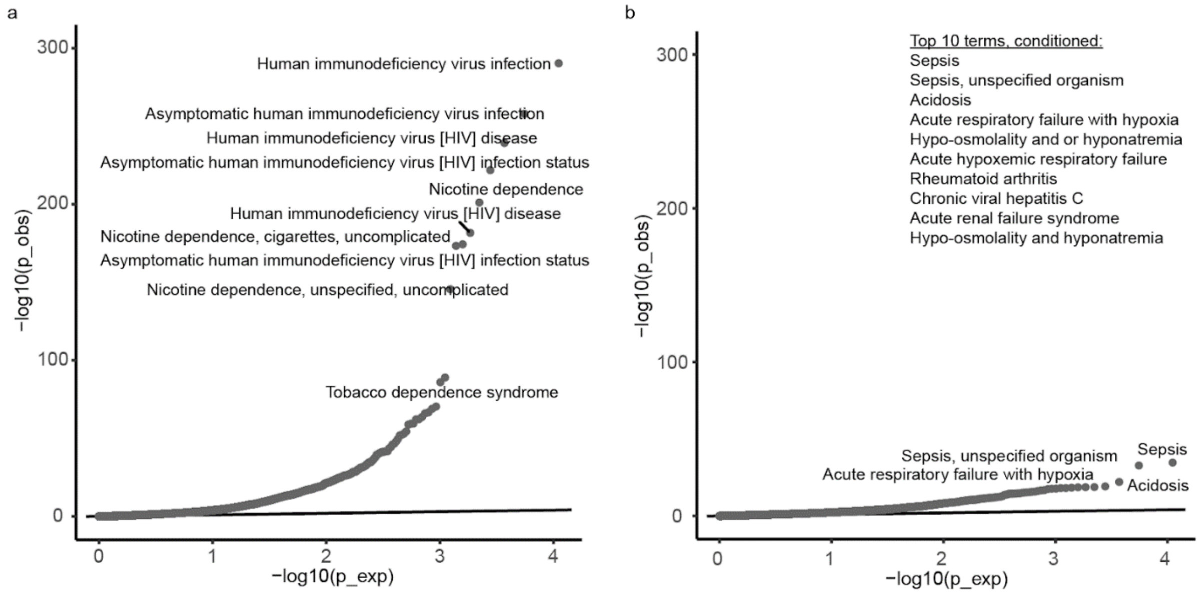


**Supplementary Figure 3: QQ-plots of the association analysis between SNOMED concept IDs and EBVread+ status within All of Us.** In the AoU-QC-cohort, associations of EBVreads with 11,111 SNOMED concept IDs were tested. **a)** The negative decadic logarithms of P-values observed for the unconditioned analysis (y-axis) are plotted against the expected distribution. In **b)** the analysis was repeated after conditioning on smoking- and HIV-status. For top10 and top4 SNOMED concept IDs, respectively, annotations are provided. All values are provided in **Supplementary Table S2**.

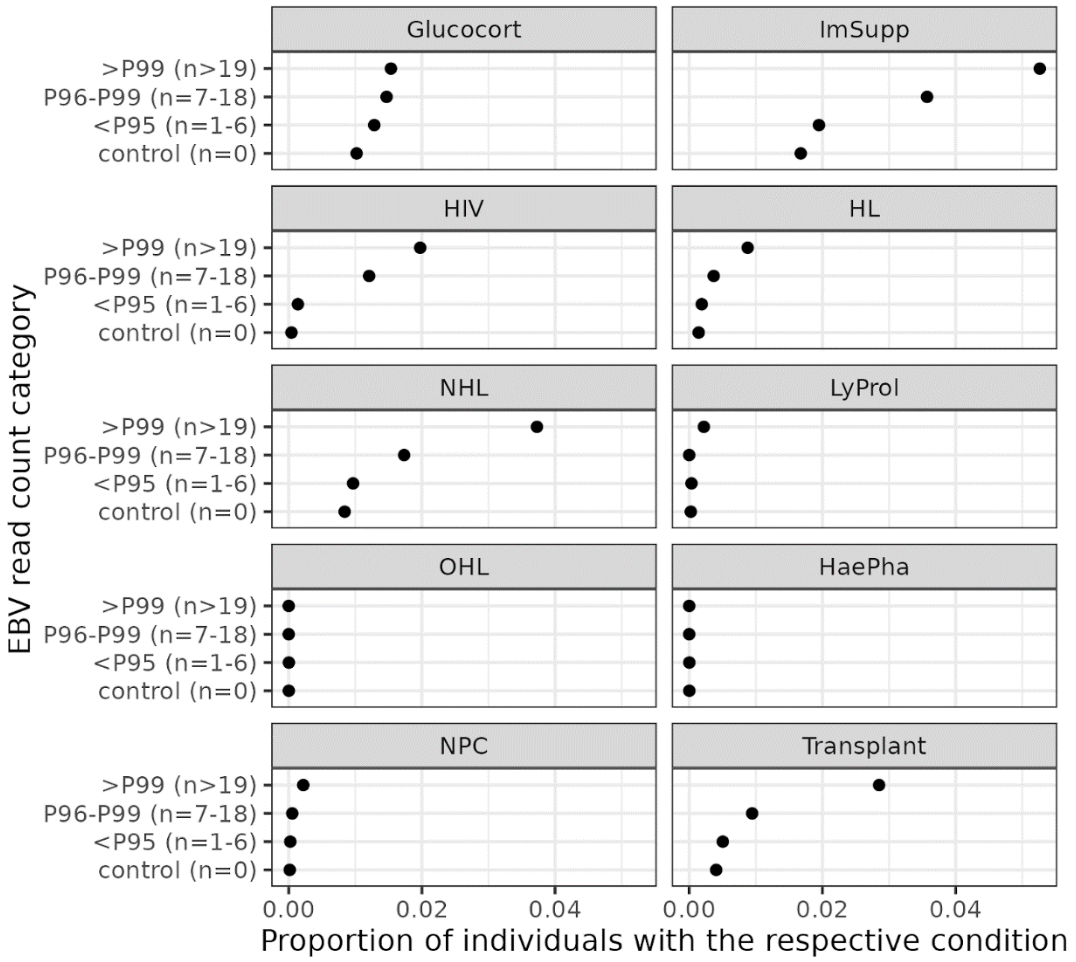


**Supplementary Figure 4: Distribution of covariates among individuals classified into different percentiles of quantitative EBV-reads**. Individuals were grouped according to their EBV read count (n) into a control group (n=0) or percentiles (P) within EBV-read-positive individuals. For each of 10 candidate conditions, data were retrieved from UKB data fields as described in **Supplementary Table S19**. This analysis shows that for the top-1% of EBV-read counts, the likelihood of some underlying malignancy increases. Therefore, individuals with 19 reads or higher were excluded from further systematic analyses. Point estimates based on n=313,843 unrelated individuals (*No-outlier-cohort* with the addition of individuals with read counts greater than 18). Abbreviations are as follows: Glucocort: Glucocorticoids; ImSupp: Immunosuppressive drugs; HIV: HIV infection; HL: Hodgkin Lymphoma, NHL: Non-Hodgkin Lymphoma; LyProl: Lymphoproliferative disorder; OHL: Oral hairy leukoplakia; HaePha: Hemophagocytic lymphohistiocytosis; NPC: Nasopharyngeal Carcinoma.

Chr. 1p13.2_PTPN22

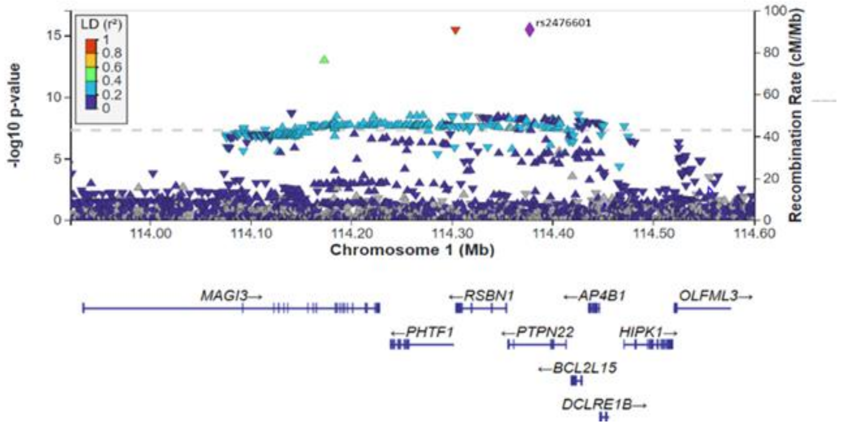


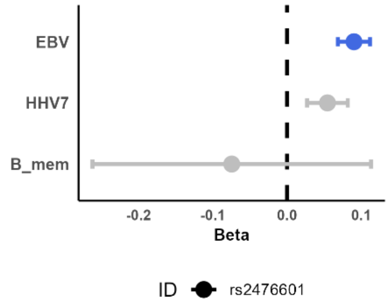


Chr. 1q23.3_SLAMF7

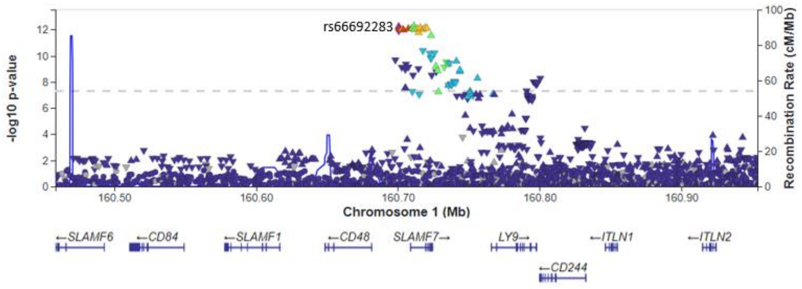


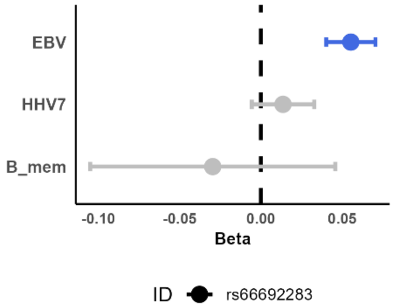


Chr. 1q25.1_PRDX6

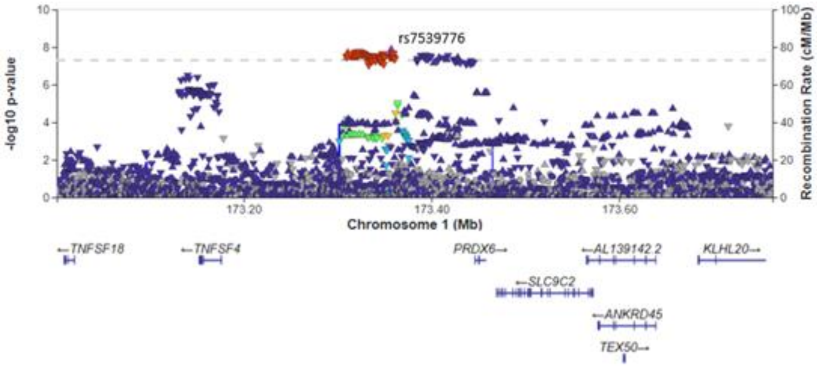


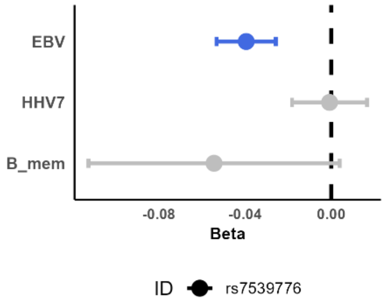


Chr. 2p22.1_SLC8A1

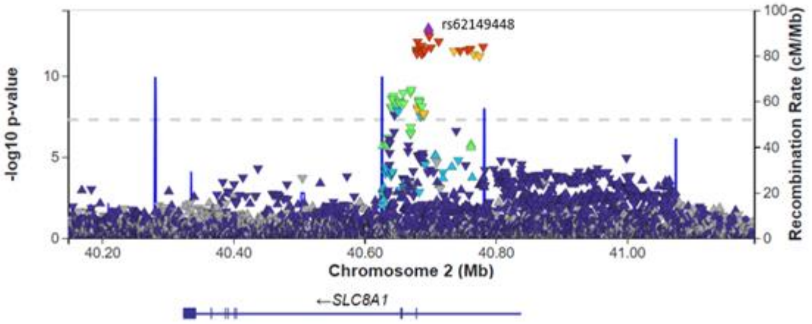


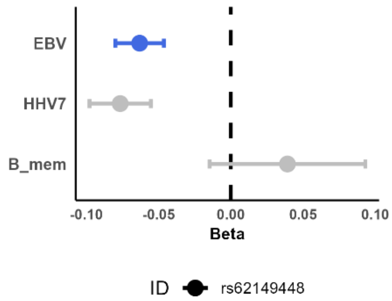

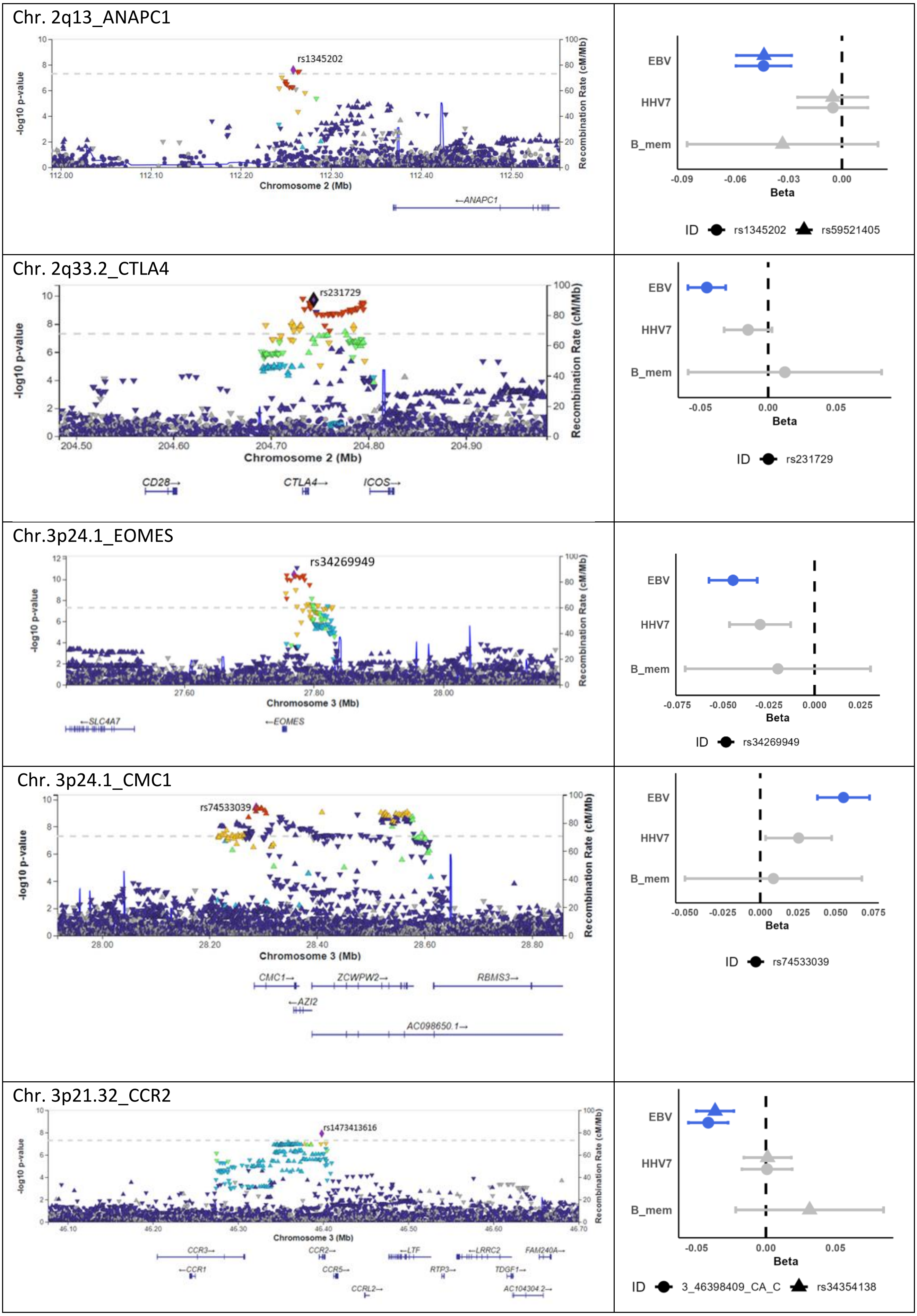
Chr. 2q13_ANAPC1
rs1345202
-log10 p-value
Recombination Rate (cM/Mb)
Chromosome 2 (Mb)
←ANAPC1
EBV
HHV7
B_mem
Beta
ID rs1345202 rs59521405
Chr. 2q33.2_CTLA4
rs231729
Chromosome 2 (Mb)
CD28→
CTLA4→
ICOS→
ID rs231729
Chr.3p24.1_EOMES
rs34269949
Chromosome 3 (Mb)
←SLC4A7
←EOMES
ID rs34269949
Chr. 3p24.1_CMC1
rs74533039
Chromosome 3 (Mb)
CMC1→
ZCWPW2→
RBMS3→
←AZI2
AC098650.1→
ID rs74533039
Chr. 3p21.32_CCR2
rs1473413616
Chromosome 3 (Mb)
CCR3→
←CCR1
CCR2→
CCR5→
CCRL2→
←LTF
RTP3→
←LRRC2
TDGF1→
FAM240A→
AC104304.2→
ID 3_46398409_CA_C rs34354138

Chr.3q13.33_ILDR1

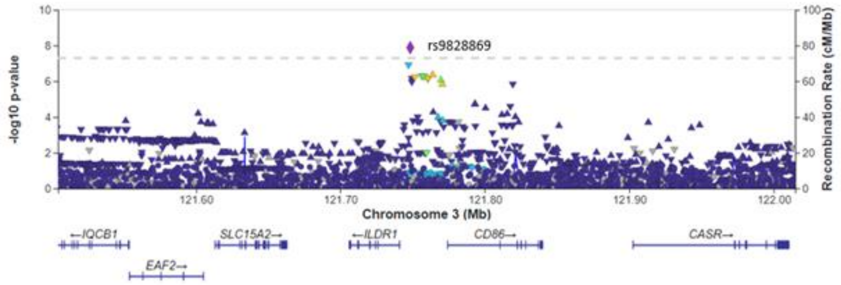


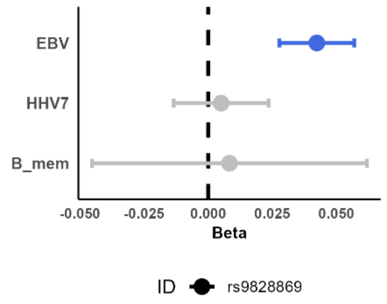


Chr. 3q21.1_PARP15

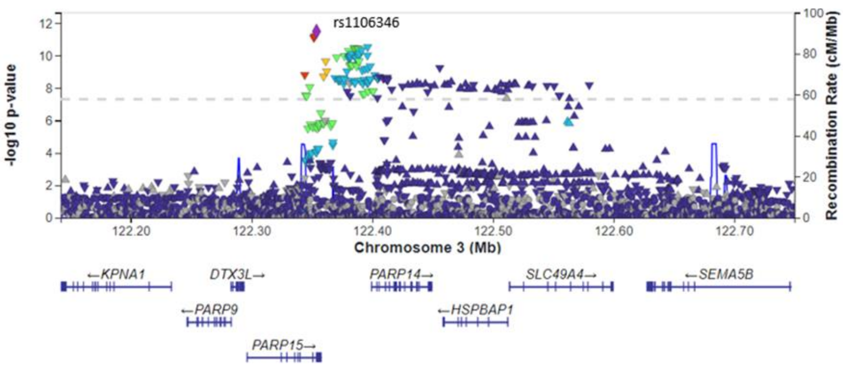


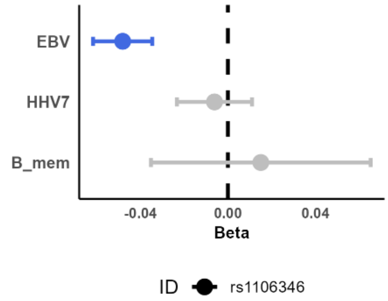


Chr. 3q28_LPP

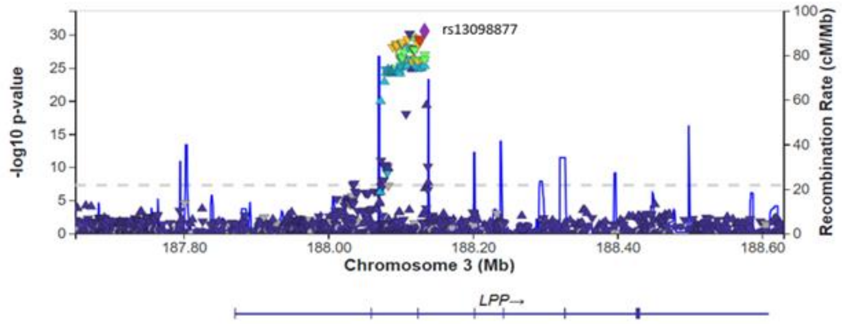


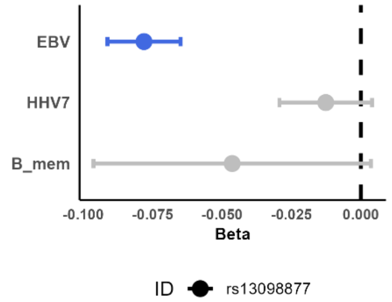


Chr. 3q28_TP63

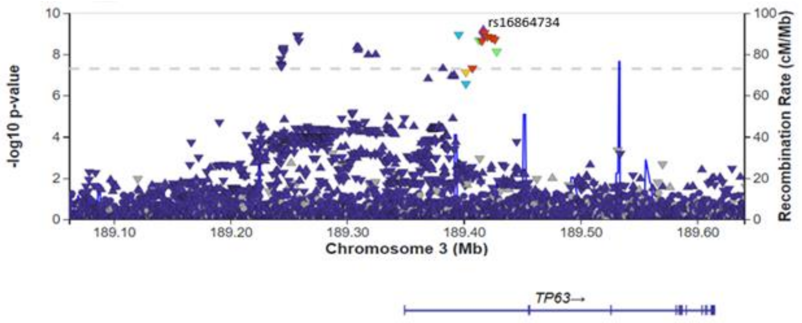


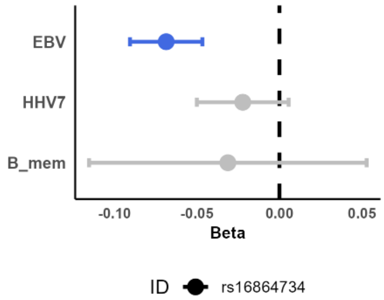

Chr. 5q15_ERAP2

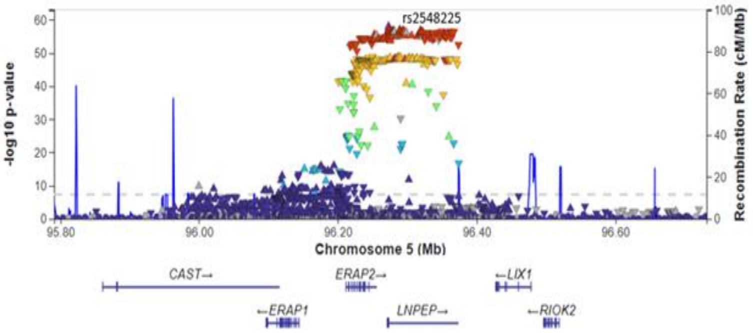


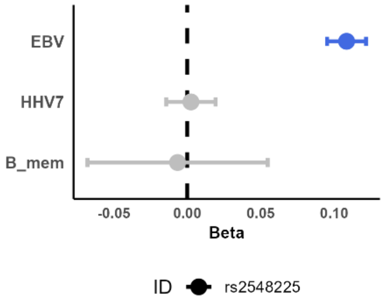


Chr. 5q31.1_IRF1

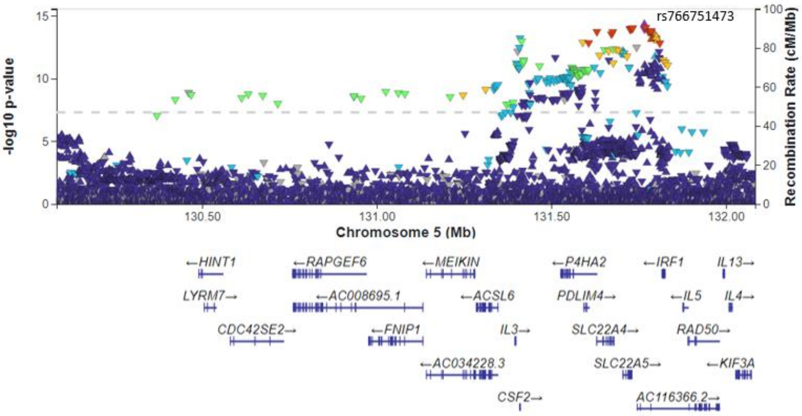


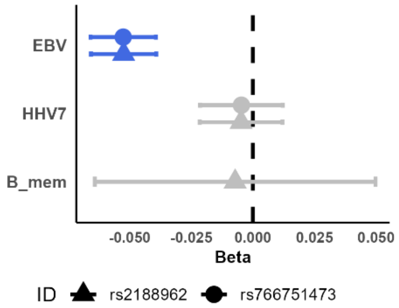


Chr. 7p12.2_SPATA48

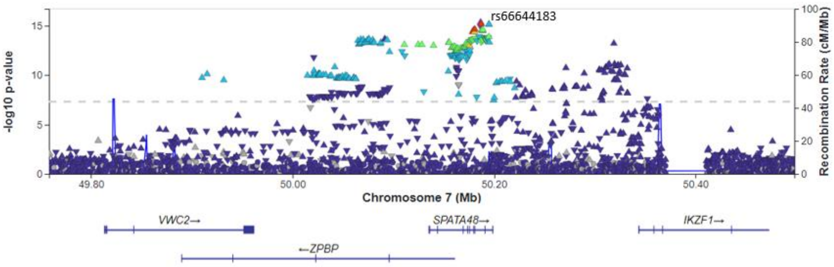


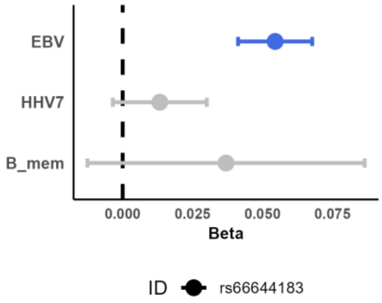


Chr. 12q24.12_SH2B3

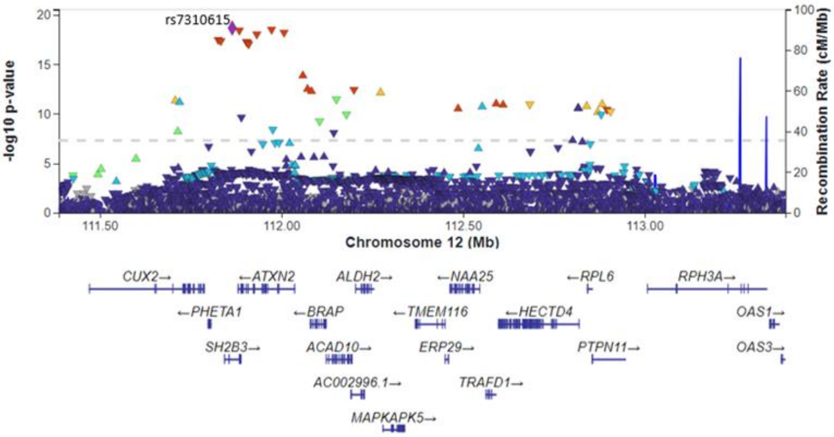


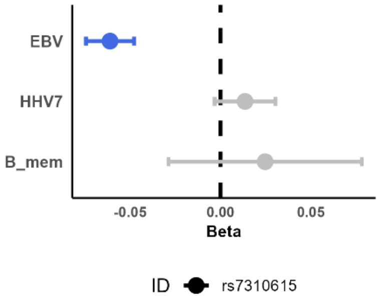

Chr. 13q14.11_FOXO1

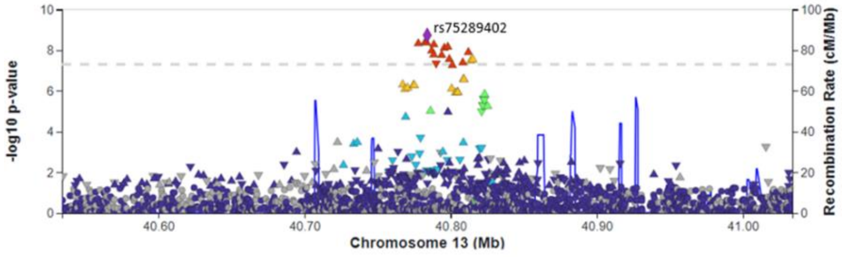


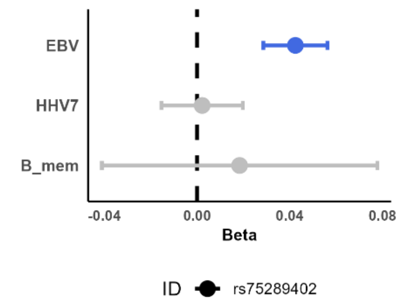


Chr. 13q32.3_UBAC2

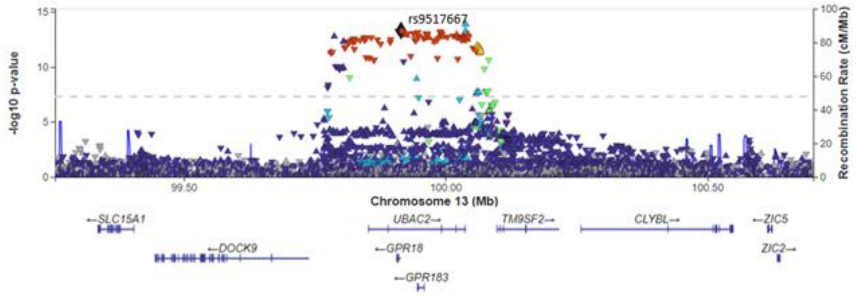


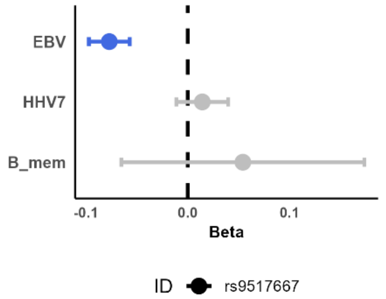


Chr. 13q33.3_TNFSF13B

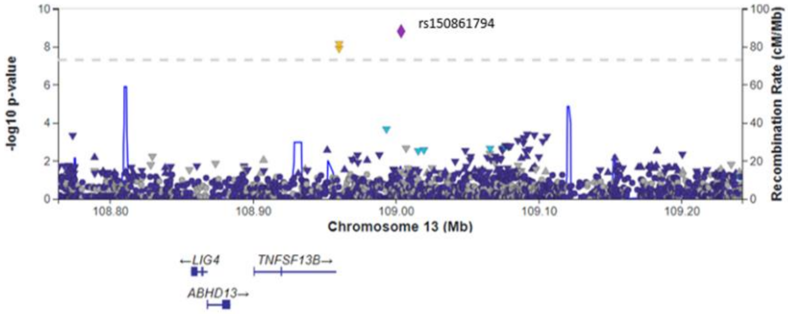


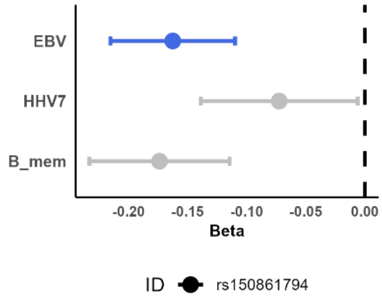


Chr. 15q15.1_CHP1

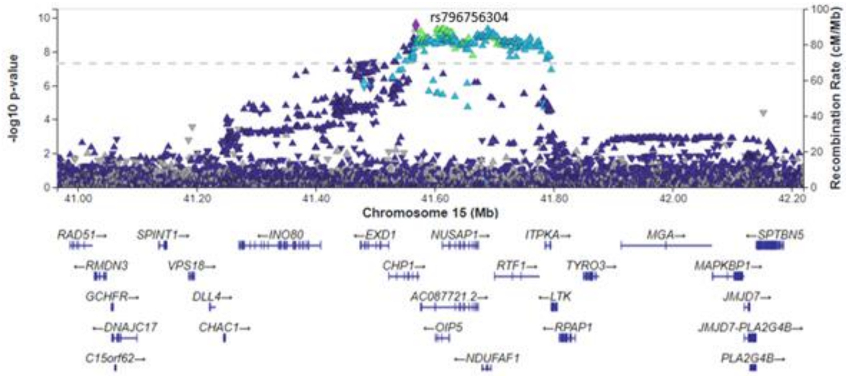


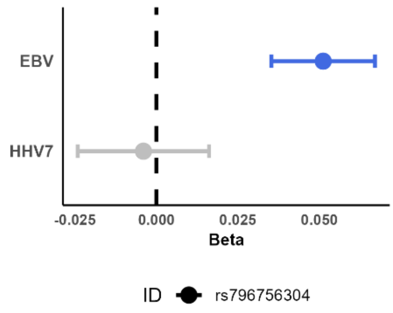

Chr. 17p11.2_TNFRSF13B

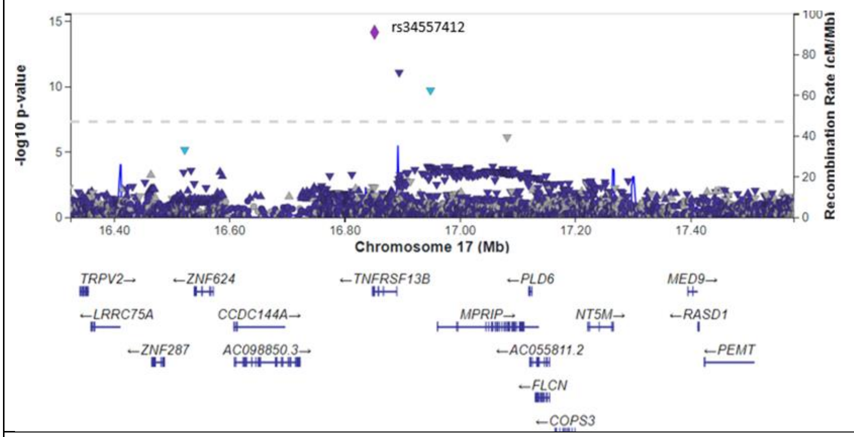


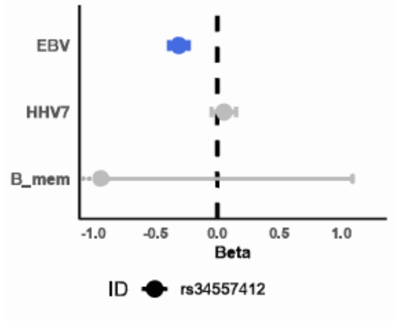


Chr. 17q11.1_KSR

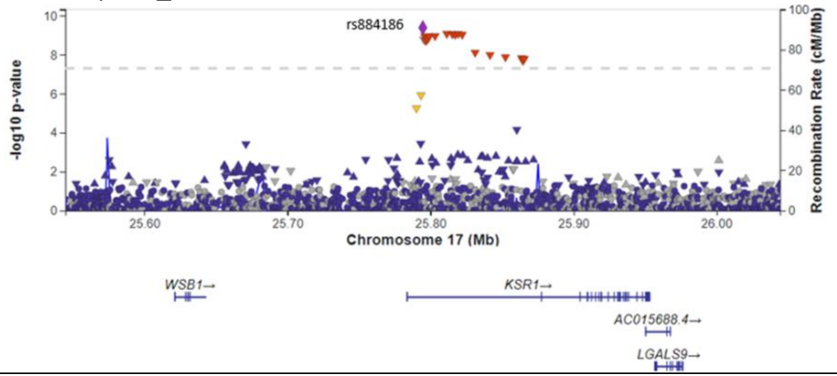


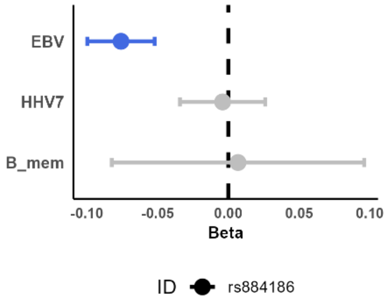


Chr. 17q12_GRB7

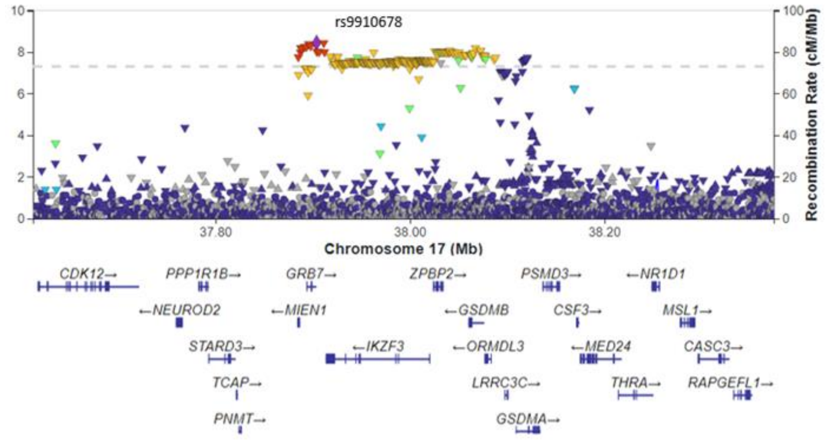


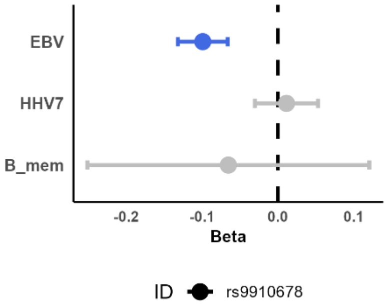


Chr. 19p13.3_CD70

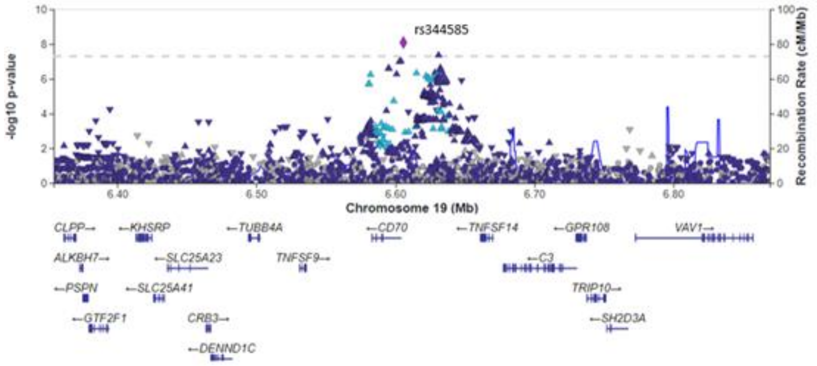


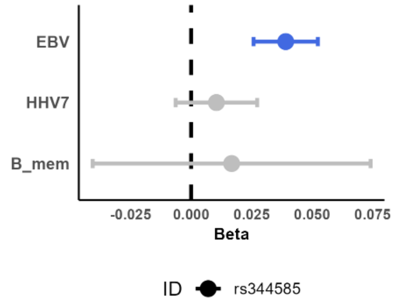

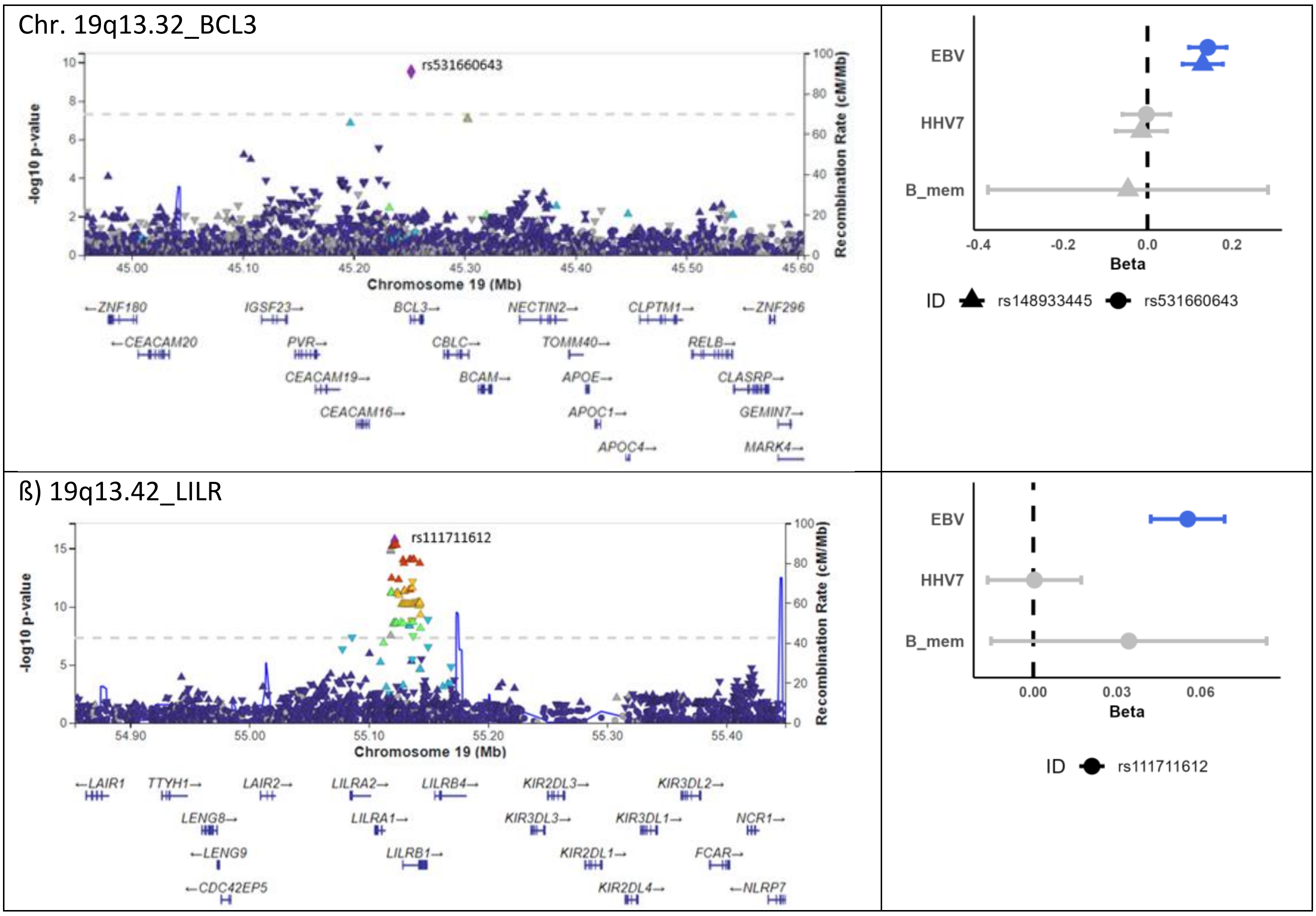


**Supplementary Figure 5: 27 loci outside the MHC region were associated at genome-wide significance with EBVread+ in UKBiobank**. For each locus, regional association plots were generated with Locus Zoom and are shown in the left column. In the right column, Forest plots provide information on the association statistics for the lead SNP of EBVread+ (unless otherwise indicated), and for HHV-7 and memory B-cells (see Methods).

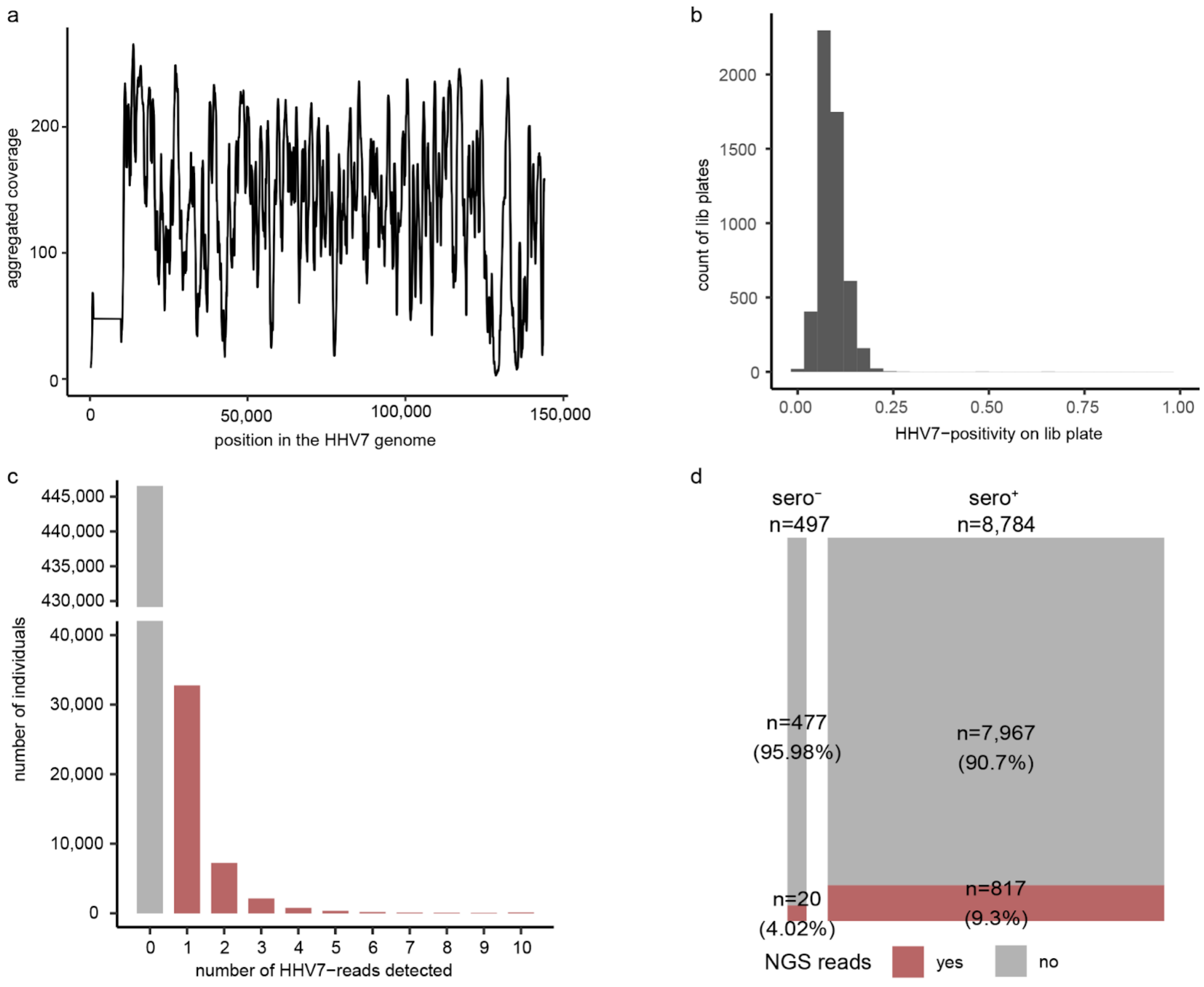


**Supplementary Figure 6: HHV-7-read extraction within UKB. a)** Cumulative read coverage across the HHV-7-genome based on the UKB-QC-cohort (line smoothed, rolling windows of 500 bp). **b)** Quality control of library plates, illustrating the distribution of HHV-7-read+ individuals per library 96-well plate. Differently from EBV, no clear indications of contaminations were observed. **c)** Number of HHV-7-reads detected per individual, with individuals showing at least one EBV-read highlighted in red (HHV-7read+). For illustration purposes, the y-axis has been interrupted between 42,000 and 430,000 individuals. **d)** Correlation of HHV-7read+ status with HHV-7 serology data (HHV-7sero-negative (sero-) and positive (sero+)), determined in the subset of the *UKB-QC*-cohort with HHV-7 serology data.

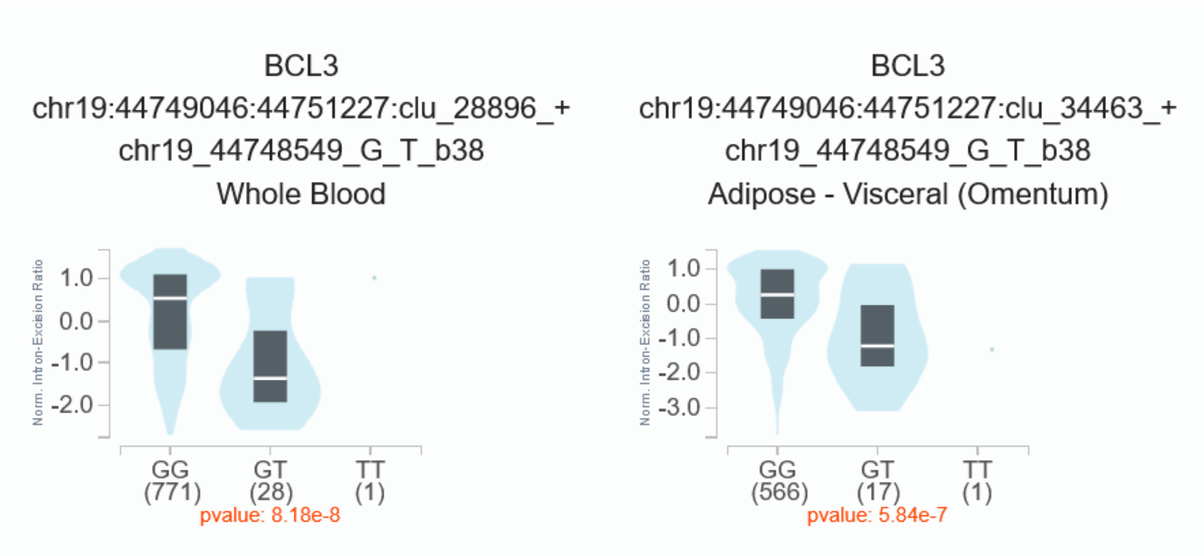


**Supplementary Figure 7: SpliceQTL effect of rs531660643 on *BCL3* in two different tissues of GTExv8.** The variant is also reported to be associated with ApoB lipoprotein levels in UK Biobank (PMID: 32203549).

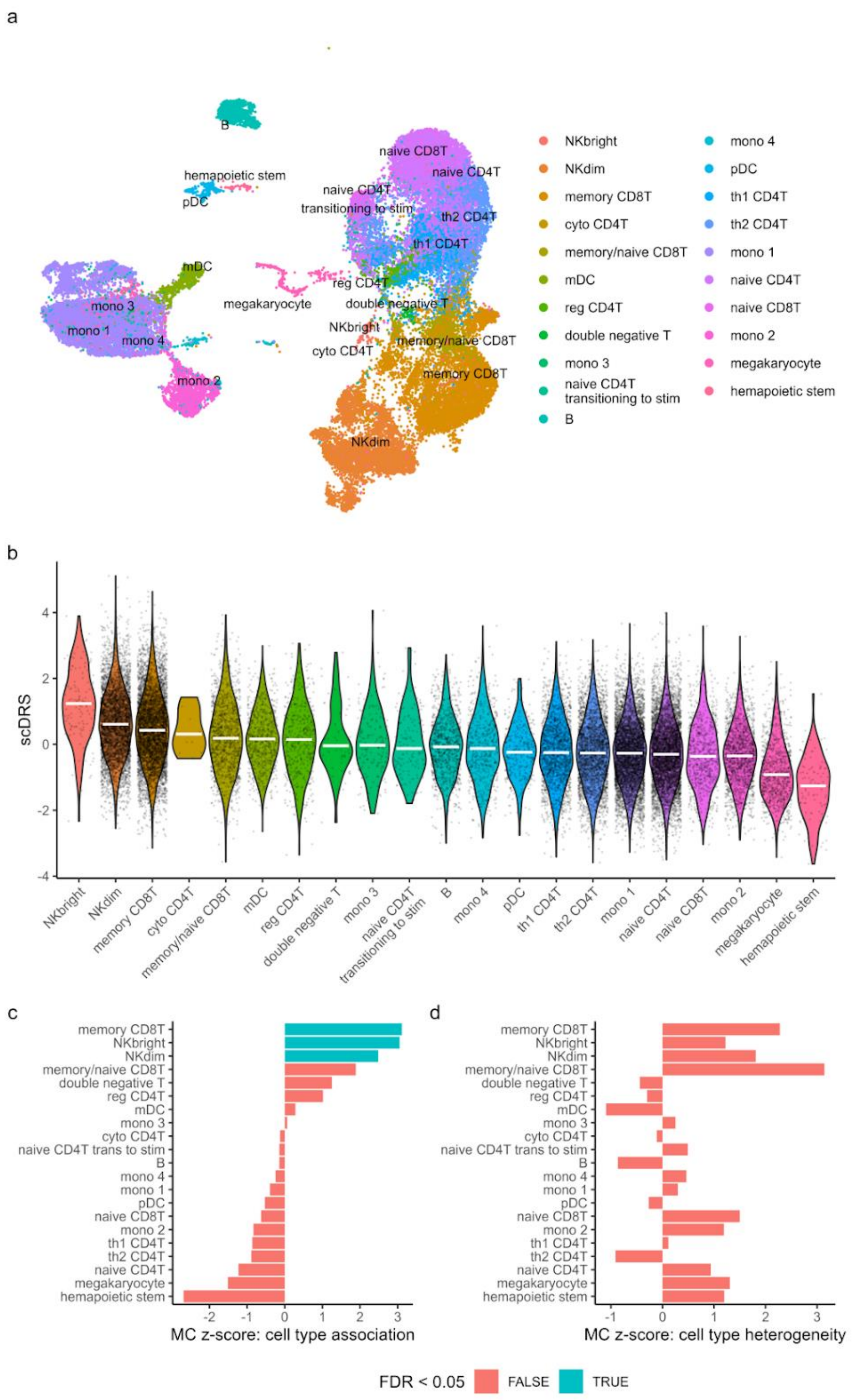


**Supplementary Figure 8: scDRS analysis as in Figure 3, using more fine-grained cell annotation. a)** UMAP representation plot of the 1M-scBloodNL data (v3) colored by cluster labels of cell type annotation level 2. **b)** Distribution of normalized single-cell disease relevance scores (scDRS) across cell types of annotation level 2, sorted by largest average score. White bars indicate the median scDRS. **c)** Results of the Monte Carlo (MC)-based statistical inference of cell type association (left) and within-cell type heterogeneity (right) with scDRS based on EBVread+. For further information, see Methods and Figure 3.

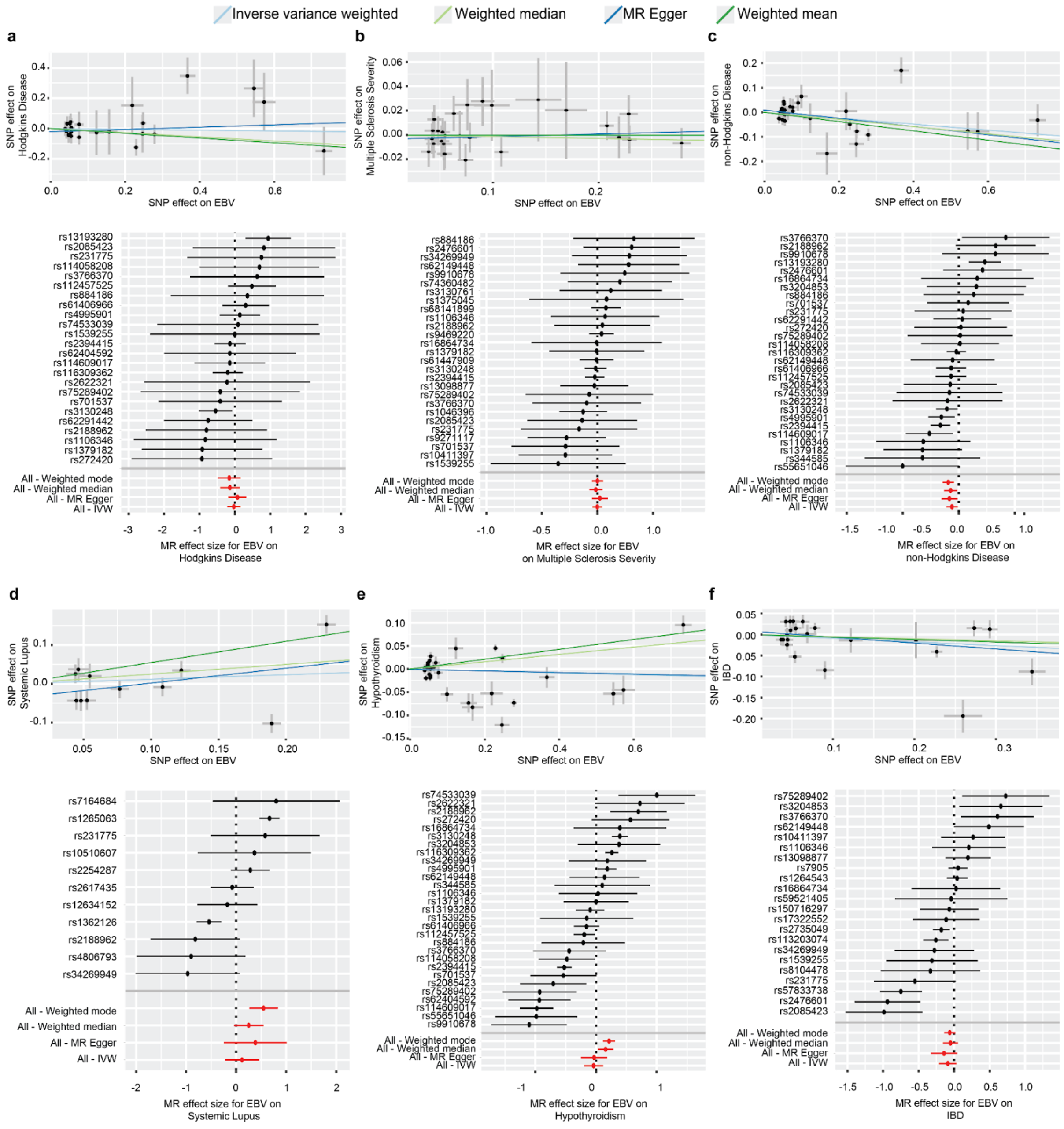


**Supplementary Figure 9: 2SMR results for the six EBV-associated diseases without evidence for causal relationships.** The figure shows scatter plots using the four standard estimators, as well as forest plots. **a)** Hodgkin's disease **b)** MS severity **c)** non-Hodgkin lymphoma **d)** Systemic lupus erythematosus **e)** Hypothyroidism **f)** Inflammatory bowel disease.

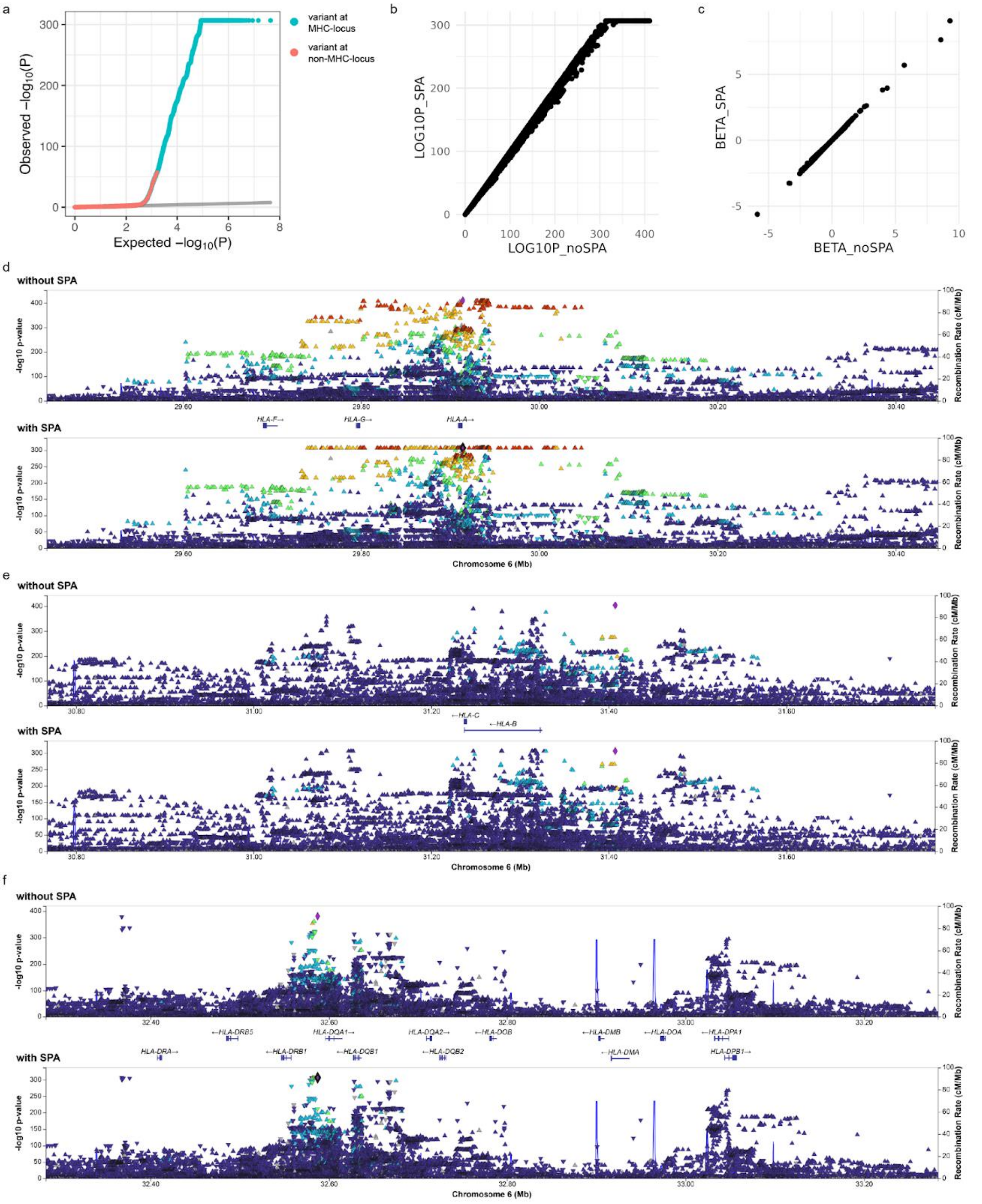


**Supplementary Figure 10: Effect of saddle-point approximation (SPA) on P-values in the MHC region. (a)** QQ-plot when conducting the GWAS using Saddle-Point-Approximation (SPA). We observed a slight inflation of test statistics (λ=1.048, LD Score regression intercept: 1.0265), which was partially attributed to the highly significant associations identified at the major histocompatibility complex (MHC) locus (without MHC region (chr6: 25-36Mb): λ=1.039). **(b)** Scatter plot of negative decadic logarithms of p-values generated by regenie with and without SPA for the MHC region (25-36 Mb). We noted that the correction of P-values by SPA that we used to counteract case-control imbalances caused a ceiling of p-values at -log10(p) of 306.653 for the most significant variants. We therefore replaced these -log10(p) values by the corresponding uncorrected values without SPA. **c)** Betas of MHC-variants with $p<0.05$ were highly correlated between the analyses with or without SPA. Panels **(d)** to **(f)** show the regional association plots for variants at the MHC region with (bottom panel) and without SPA (top panel), respectively, divided into regions approx. 1 Mb. In between the panels, HLA-genes are shown, respectively.
