## Supplementary Notes for "Host control of latent Epstein-Barr virus infection"

**Supplementary Note 1:**

Code snippets to extract and filter EBV and other viral reads are given below. Note that the file path to the CRAM-file (CRAM_FILE_PATH) needs to be set and a fasta file with viral genome sequences (VIRUS_REFERENCE_SEQS.fasta) needs to be provided. The program versions used were 2.2.1 for bwa-mem2 and 1.20 for samtools.

EBV-reads (based on reference genome available in GRCh38):

```
samtools view -e "rnext==rname" -h "$CRAM_FILE_PATH" chrEBV >
raw_reads.sam
```

```
samtools view raw_reads.sam -e 'sclen<=20 && rlen>=120' -F 1024 -O
BAM -o filtered_reads.bam
```

```
samtools view filtered_reads.bam | cut -f1,4 >
reads_mapping_to_EBV.tsv
```

Other viral reads (no reference sequence available in GRCh38; used for HHV-7):

```
bwa-mem2 index VIRUS_REFERENCE_SEQS.fasta
```

```
samtools view -f 12 -u "$CRAM_FILE_PATH" "*" | samtools fastq -1
FASTQ_FILE_1.fq.gz -2 FASTQ_FILE_2.fq.gz -0 /dev/null -s /dev/null -
n
```

```
bwa-mem2 mem VIRUS_REFERENCE_SEQS.fasta FASTQ_FILE_1.fq.gz
FASTQ_FILE_2.fq.gz | samtools view -F 12 -h > VIRUS_READS.sam
```

```
samtools view VIRUS_READS.sam -h -q 1 -e 'sclen<=10 && rlen>=130 &&
rnext==rname' -F 1024 | samtools view -f1 -O BAM -o
VIRUS_READS_FILTERED.bam
```

```
samtools view VIRUS_READS_FILTERED.bam | cut -f1,3,4 >
reads_mapping_to_viruses.tsv
```

**Supplementary Note 2:**

The robustness of our results suggest that a GS coverage of 30x is sufficient to identify those individuals that control EBV better than others, despite a technical detection limit. Assuming that all sequenced cells would carry one copy of EBV, one would expect N reads mapping to EBV:

N=G * CL = 172,000bp * 152x150bp =8,600
Where G corresponds to the genome size of EBV, C the sequencing coverage attributed to a haploid genome and L the read length. Therefore, if 8,600 reads correspond to all cells carrying one copy of EBV, one read corresponds to one EBV genome in 8,600 cells.

When we consider that EBV mainly resides in memory B cells, which make up ~0.6% of leukocytes (29476184), detection of a single EBV-read would correspond to approximately 1 EBV genome among 55 memory B cells.

In a study using qPCR the prevalence of EBV in PBMCs was given to be 11.03% (PMID 26744460). This study stated a detection limit of one EBV genome in 2,000 PBMCs, with PBMCs generally making up about 30-40% of the total leucocyte count. Following from the calculation given above, the presence of one EBV read would translate into about one EBV genome in 2,580 to 3,440 PBMCs. From these numbers, our GS-based approach for detecting EBV is expected to be slightly more sensitive than the qPCR-based approach, which can explain the comparable, but slightly higher prevalence of EBV-reads we have identified.

**Supplementary Note 3:**
To check the robustness of positive 2SMR findings, we performed a range of additional sensitivity analyses. Some effects were sensitive to leave-one-out analyses (Supplementary Table S23). Analyses that suggested a causal effect using two or more estimators (after Bonferroni-correction) were followed up using the outlier- and pleiotropy robust estimators MR-RAPS and MR-PRESSO, and we further examined the individual SNPs driving these effects. For example, using 14 SNPs as instruments, 2SMR analyses suggested a potential causal effect of EBVread+ on MS using two out of four estimators (ORwMod = 1.57, 95% CI = [1.37, 1.81], ORwMed = 1.53, 95% CI = [1.24, 1.89]), which was however, not robust to several sensitivity analyses and seemed to be driven by SNPs located in the MHC region (Supplementary Table S22 and Extended Data Figure 4). While there was no evidence for horizontal pleiotropy as indicated by the MR-Egger intercept (Int = –0.005, s.e. = 0.03, P = 0.870), we observed significant heterogeneity of effects (Q = 148.65, df = 12, P = $1.07 \times 10^{-25}$). Furthermore, MR-PRESSO showed significant outlier effects, and the causal effect estimates both before and after outlier exclusion were non-significant when using MR-PRESSO (OR= 0.94, 95% CI = [0.68, 1.32] and OR = 0.79, 95% CI = [0.59, 1.07], respectively). Results for rheumatoid arthritis (RA) and type 1 diabetes (T1D) were similar but somewhat more robust, with significant effects across all estimators, including the robust estimators MR-RAPS (OR = 1.67, 95% CI = [1.47, 1.91] and OR = 1.90, 95% CI = [1.72, 2.11], for RA and T1D, respectively) and MR-PRESSO (OR = 1.34, 95% CI = [1.04, 1.72] and OR = 1.62, 95% CI = [1.31, 2.00], respectively). Notably, these pleiotropy robust estimators showed significant effects despite the presence of significant horizontal pleiotropy for the T1D analysis (MR-Egger intercept = –0.04, *s.e.* = 0.001, *P* = 0.001). However, all these effects also attenuated towards null when restricting to non-MHC SNPs (e.g., RA: $OR_{wMed}$ = 1.06, 95% CI = [0.84, 1.34]; T1D: $OR_{wMed}$ = 0.96, 95% CI = [0.67, 1.40]). The negative control trait (hair colour: red) did not show any significant effects (e.g., $OR_{wMed}$ = 1.02, 95% CI = [0.97, 1.08]).
